# Intratumor bacteria is associated with prognosis in clear-cell renal cell carcinoma

**DOI:** 10.1101/2023.12.29.23300629

**Authors:** Yuqing Li, Dengwei Zhang, Linyi Tan, Junyao Xu, Ting Guo, Yang Sun, Rui Zhang, Yao Cheng, Haowen Jiang, Wei Zhai, Yong-xin Li, Chenchen Feng

## Abstract

**Background:** Intratumor bacteria (ITB) plays a role in various cancer types. Its role in clear-cell renal cell carcinoma (ccRCC) remains elusive due to small sample size and inadequate decontamination in relevant studies.

**Objective:** To establish common and reproducible ITB-associated biomarkers in ccRCC.

**Design, setting, and participants:** This retrospective study comprised seven bulk RNA sequencing datasets from six publicly available cohorts and one in-house Chinese cohort (Renji), one 16S rRNA sequencing dataset from an original Chinese cohort (Huashan), and one publicly available single-cell RNA sequencing dataset. All of these datasets included ccRCC cases.

**Outcome measurements and statistical analysis:** Composition was presented by relative abundance. Overall and progression-free survival were primary outcomes profiled by putative ITB load and risk score, respectively. Potential host interaction was exploratorily analyzed using gene set enrichment analysis and Sparse CCA.

**Results and limitations:** Nine cohorts encompassing a total of 1049 ccRCC cases and 130 paired normal tissues were initially analyzed and underwent decontamination. Surprisingly, neither diversity nor composition was differentially distributed between normal and cancer tissue. High putative bacterial load was associated with better overall survival. Notably, a 7-genera dichotomized ITB risk score was indicative of overall survival and a 13-genera dichotomized ITB risk score was predictive of progression-free survival, respectively. *Actinomyces*, *Rothia* and *Bifidobacterium* showed a protective role while *Exiguobacterium* was a risk factor. A limitation is lack of causation analyses.

**Conclusions:** ITB exists in ccRCC. High ITB loads and ITB-risk score predicts better ccRCC survival regardless of sequencing tech, sample processing or racial disparity.

**Patient Summary:** In this report, we explored the role of intratumor bacteria (ITB) in renal clear-cell carcinoma (ccRCC) in patients with different race and sequencing platforms. Putative ITB load and a 7-genera ITB risk score were associated with overall survival. A 13-genera ITB risk score was predictive of progression-free survival. We conclude that certain ITB features are universally pathogenic to ccRCC.

## INTRODUCTION

Intratumor microorganisms, especially intratumor bacteria (ITB) signature has shown prognostic effect in a variety of cancer types including gastric cancer^1^, colorectal cancer^2^, hepatocellular cancer^3^, etc., establishing microbiome as a novel omics or “second genome” of cancer^4^. However, ITB may vary to vast extents that renders intratumor microbe findings in some cases, hardly reproducible^5^. Amongst all confounders, biomass of subject^6,7^, race^8,9^, sequencing tech^10^., and contamination^11^ play the most pivotal roles.

Despite the pitfalls present in ITB studies, true bacterial signatures could be still identified by imperfect sequencing technologies and decontamination processes, which has been demonstrated through experimental validation^12,13^. Furthermore, when consistent findings emerge from multiple cohorts, the influence of these pitfalls can be minimized, leading to more reliable and robust conclusions.

Clear-cell renal cell carcinoma (ccRCC) is the most common type of malignancy in kidney that is conventionally accepted as sterile organ and ccRCC is expected to harbor a low biomass of ITB. To date, the existing literature on ITB in ccRCC is limited to three full papers^14–16^ and one meeting proceeding^17^. These studies suffer from small sample sizes, lack of racial diversity, use of a single sequencing technology, and inadequate decontamination process. Consequently, the precise composition and significance of ITB in ccRCC remain elusive. Therefore, a multi-cohort study focusing on ccRCC is urgently required.

While we highly concur that ITB exists in most, if not all solid tumors including ccRCC, we aim to answer whether common ITB composition exists in ccRCC and whether ITB signature is prognostic, regardless of demographic, racial and sequencing differences. To achieve this, we incorporated various reports on decontamination and multiple ccRCC cohorts encompassing over 1000 cases that vary in region, race, batch, sequencing tech, etc. We aim to identify inherent ITB signatures and explore its prognostication in ccRCC in the current study.

## METHODS

### Study Population

For 16S rRNA sequencing, we retrospectively collected the 217 tumor and 27 normal samples from 217 patients histologically diagnosed clear cell renal carcinoma who underwent partial or radical nephrectomy in Huashan hospital (Shanghai, China) between May 2013 and Oct 2022 under reasonable inclusion criteria (**Figure 1**). The samples were formalin-fixed and paraffin-embedded. We also included 10 negative controls using sliced paraffin from the margin of the block, sampling paraffin only without tissue. Tumor stages were stratified according to the 8th American Joint Committee on Cancer staging system (AJCC)^18^ No subjects received preoperative treatments, including immunotherapies or molecular targeted therapies.

**Figure 1.**
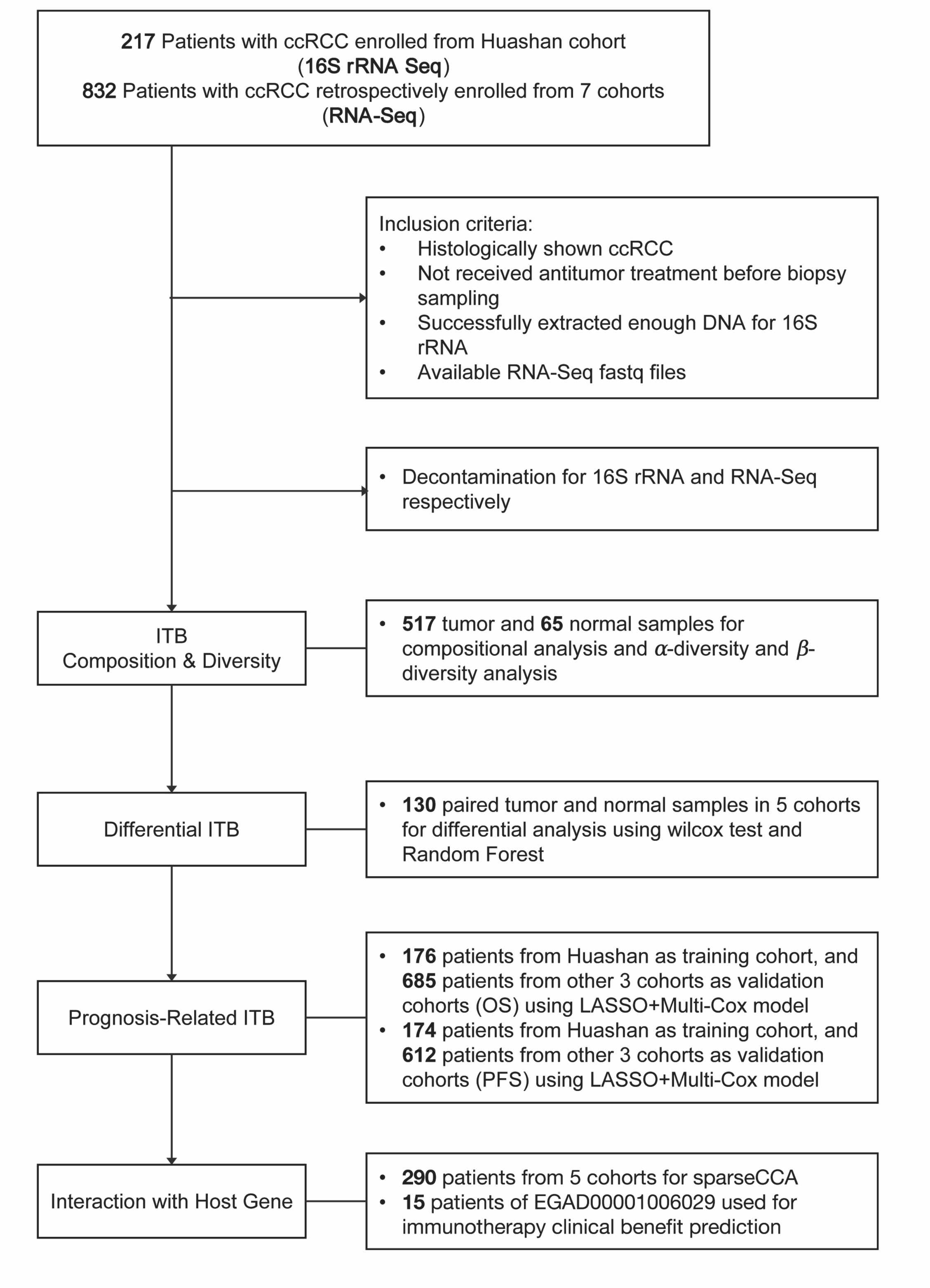
Study Design.

Six cohorts with the RNA-Seq data available were included in our study. EGAD00001000597^19^ as an integrated molecular study of ccRCC and consists of 100 tumor samples. EGAD00001006029 (CheckMate 025; NCT01668784)^20^ was a prospective clinical trials of the anti-PD-1 antibody nivolumab in advanced clear cell renal cell carcinoma, and 53 FFPE tumor tissues were obtained prior to initial therapy for patients enrolled in this study, including 15 patients with the objective response record of immunotherapy. Data from above two datasets (EGAD00001000597 and EGAD00001006029) were requested from the principal strictly via European Genome-phenome Archive (EGA, https://ega-archive.org/) according to the clinical data transfer agreement. GSE102101^21^, GSE126964^22^, GSE151419^23^ were studies concerning on the renal cell carcinoma by organizations located in Singapore, China and Poland, respectively, and the raw sequencing data of tumor and paired normal samples were downloaded from the gene expression omnibus (GEO) data repository (https://www.ncbi.nlm.nih.gov/geo/). Besides, 27 fresh tumor samples with RNA sequencing were supplied by the Renji hospital (Shanghai, China).

TCGA was a cancer genomics program which molecularly characterized primary cancer and matched normal samples including clear cell renal cell carcinoma (KIRC). However, due to the limit of access to the level 1 or 2 data of TCGA hosted at the Genome Data Commons (GDC) website, the microbiome data processed by by Poore et al. ^24^ and by Salzberg et al. ^25^ respectively were directly adopted for downstream bioinformatic analysis in this study. Poore et al. derived the microbiome data from both WGS and RNA-Seq data, and we used the normalized and batch effect-corrected data of 532 tumor and 72 paired normal samples. The author performed decontamination in several degrees and got 5 microbial communities including data with non-contamination removed (NR), data with likely contaminants removed (LR), data with putative contaminants removed (PR), data with contaminants removed by sequencing “plate–center” combinations (CR), and data with mostly stringent filtering (SR). Salzberg et al. also used the TCGA data but only took WGS data into consideration and shared us with the data including 40 tumor and 35 paired normal samples. Demographics of all cohorts were demonstrated in **Table 1**.

### 16S rRNA gene amplicon sequencing

In preparation for the 16S rRNA gene sequencing, samples were sectioned from the paraffin-embedded tissue blocks, which accepted quality testing, purification and nested amplification. To meet the requirement of sufficient DNA for sequencing, the amplified products were detected by DNA electrophoresis, and the eligible samples were kept for further study. 16S rRNA gene sequencing was conducted at the Nonogene Co., Ltd. In brief, genomic DNA was extracted from the tissue samples using the CTAB/SDS method. The 16S rDNA V4 region was amplified through PCR employing a primer pair (515F: 5’-GTGCCAGCMGCCGCGGTAA-3’ and 806R: 5’-GGACTACHVGGGTWTCTAAT-3’) with a barcode. Sequencing libraries were prepared using the TruSeq® DNA PCR-Free Sample Preparation Kit (Illumina, USA) following the manufacturer’s instructions. The libraries were subsequently sequenced on the Illumina NovaSeq platform, yielding 250 bp paired-end reads.

### 16S rRNA sequencing data processing and analysis

The raw sequencing data of 16S rRNA in FASTQ format underwent processing with QIIME 2 version 2023.2. Quality filtering, denoising, and chimera removal were performed using DADA2, resulting in high-quality sequences that were assigned to amplicon sequence variants (ASVs). The feature table was constructed using these ASVs, and taxonomic information was annotated using a Naive Bayes classifier trained with the SILVA 138 SSURef NR99 database. ASVs that could not be confidently assigned at the phylum level, as well as non-bacterial ASVs, were excluded from further analysis.

To minimize the potential impact of contaminants, we employed a previously established decontamination process. This involved three steps: Firstly, we used the isNotContaminant function in the "decontam" algorithm (ref) to identify possible contaminants. This prediction was based on the difference in ASV prevalence between FFPE samples and tissue samples. Secondly, ASVs with a relative abundance greater than 0.5% in the FFPE samples were removed. Lastly, ASVs that appeared in less than 5% of the tissue samples were further eliminated to avoid contingency. Only ASVs that met these criteria were retained for downstream analysis.

### Bulk RNA sequencing data processing

Raw RNA sequencing data from tissue samples obtained from six cohorts were acquired online. Sequencing reads were quality-controlled using fastp v0.21.1, with parameters “-l 50 -5 3 -3 3”. Filtered reads that were shorter than 50 bp were discarded. To quantify human gene expression, the clean reads were aligned to the human reference genome, GRCh38.p13, available in the GENCODE database using HISAT2 v2.2.1. The gene expression values were quantified in transcripts per million (TPM) using StringTie v2.2.1.

For profiling the intratumor bacteria from bulk RNA-Seq data, clean reads were initially aligned against an indexed database to remove host or contaminant reads. This alignment was performed using bowtie2 v 2.4.5 with a “--very-sensitive” model. The indexed database included 9 mammalian genomes (hg38, felCat9, canFam6, mm39, rn7, rheMac10, susScr11, galGal6, bosTau9; University of California– Santa Cruz Genome Browser), 2145438 complete bacterial plasmids (PLSDB databse, v.2021_06_23_v2), 13705 mitochondrial genomes (NCBI RefSeq database, accessed on Aug 15, 2022), 9443 plastid sequences (NCBI RefSeq database, accessed on Aug 15, 2022), 6093 UniVec sequences (NCBI RefSeq database, accessed on Aug 15, 2022), which were considered potential sources of human habitat- or laboratory-associated or extrachromosomal sequence contaminants for taxonomic classification of microbial metagenomic sequences^26^. Unmapped paired reads were then subjected to KrakenUniq v 1.0.4 for taxonomic assignment using a pre-built database. This database includes complete microbial genomes from RefSeq, comprising 46,711 bacterial genomes, 13,011 viral genomes, and 604 archaeal genomes. Additionally, the database contains 246 eukaryotic pathogens, the UniVec set of standard laboratory vectors, and the GRCh38 human genome. The abundance of bacteria was evaluated at the genus level, which was deemed more accurate than the species level.

To ensure the removal of potential false positive assignments, the bacterial genera underwent further filtration based on the following criteria: (1) the genus must contain more than 5 reads; (2) number of duplicated kmer must be larger than half of assigned read counts; (3) genome coverage must be larger than 1e-5. (1) the genus must have a read count greater than 5; (2) the number of duplicated k-mers must exceed half of the assigned read counts; and (3) the genome coverage must be larger than 1e-5. Additionally, efforts were made to distinguish the potential host of the identified genera in order to eliminate non-human-associated genera that are likely to be contaminants. To accomplish this, information regarding the isolation sources of bacteria deposited in NCBI (https://www.ncbi.nlm.nih.gov/genome/browse#!/prokaryotes/), IMG/M (https://img.jgi.doe.gov/cgi-bin/m/main.cgi), GOLD (https://gold.jgi.doe.gov/downloads), and BV-BRC (https://www.bv-brc.org/docs/quick_references/ftp.html) was gathered. For identified genera not isolated in these four databases, potential hosts were obtained through a literature search on Google Scholar. Based on the available host information, the identified genera were classified into three groups: non-human (genera not isolated from human), human-exclusive (genera exclusively associated with the human host), and mixed (genera derived from either human or other environments). Non-human-associated genera were subsequently excluded from further downstream analysis.

### Microbial analysis

For 16S rRNA sequencing data, feature table, taxonomy, and phylogenetic tree after decontamination were combined into a Phyloseq object for downstream processing. To estimate alpha diversity and beta diversity, all samples were rarefied to 2000 sequencing reads. The statistical significance of differences in alpha diversity was assessed by stat_compare_means function in R package “ggpubr”. Difference in microbial compositions was tested using Permutational multivariate analysis of variance (PERMANOVA).

For the bulk RNA-Seq data, the counts of genera were converted to relative abundance for analysis. Due to the ununiform sequencing depth that would skew the measure of alpha diversity, we did not examine alpha diversity among RNA-Seq data. Rather, we compared the bacterial read counts per million reads, which could provide an indication of bacterial load. The Bray-Curtis dissimilarity among the samples was calculated using the vegdist function in the R package "vegan" and subjected to Principal Coordinates Analysis (PCoA). The statistical significance of the findings was evaluated using PERMANOVA analysis with the adonis2 function.

To evaluate the impact of clinical factors on intratumor microbial communities, a PERMANOVA analysis with 999 permutations was conducted based on Bray-Curtis dissimilarity. To account for multiple comparisons, all P-values were adjusted using the false discovery rate (FDR) method. To explore the relationship between the overall microbial community and overall survival or progression-free survival, dimensionality reduction was employed to reduce the complexity of the microbial data. Principal Component Analysis (PCA) was performed using the PCA function in the "FactoMineR" R package. The first five principal components (PCs) of the intratumor microbiome PCA were retained to represent the overall intratumor microbiome. Cox proportional hazard regression models were employed to examine the association between each PC and overall survival or progression-free survival. This analysis was conducted using the coxph function in the "survival" package. P-values were adjusted for multiple comparisons using FDR methods.

### Identifying diagnosis-related microbiome

For differential abundance testing between tumour and normal tissues in ccRCC, we used relative abundance and counts per million reads (CPM) respectively. We performed Wilcoxon rank-sum tests for each feature in genus level, and corrected the resulting p-values with the BH method. To exclude the bias caused by the sample number imbalance, we incorporated only the matched specimen and finally got 24 pairs in Huashan cohort, 10 pairs in GSE102101 (Cohort 3), 11 pairs in GSE126964 (Cohort 4), 13 pairs in GSE151419 (Cohort 5).

We also used the Random Forest algorithm to further identify the potential features distinguishing the paired samples using randomForest function in the R package " randomForest"^27^. Ten-fold cross-validation and five repetitions were adopted to help select a specific number of features, whose importance were measured by accuracy and Gini index.

### Identifying prognosis-related microbiome

Difference in microbial compositions was first tested between population with long term survival (LTS) and short term survival (STS). Due to the inconsistent following months, we used the median survival time in each cohort as the cutoff. Permutational multivariate analysis of variance (PERMANOVA) was used for testing difference in microbial compositions as above mentioned.

Univariate cox was performed to identify the genera whose abundance associated with overall survival and progression free survival. The HR (Hazard Ratio)>1 indicated that the feature was a risk factor for the prognosis, while HR<1 indicated that the protect factor. A cluster of genera were preliminarily screened as the input for the least absolute shrinkage and selection operator (LASSO) to exclude the features with potential multi-collinearity. The glmnet function in the R package "glmnet"^28^ was used and the family was set as "cox" while the other parameters were set default. Finally, we constructed the cox model using coxph function in the R package " survival". To fit the model more reasonably, we took the stepwise regression method to help select a formula-based model by Akaike information criterion (AIC). The OS-related risk cox model consist of 7 genera, including Abiotrophia, Actinomyces, Bifidobacterium, Dolosigranulum, Faecalibacterium, Kocuria, and Prevotella. The PFS-related risk cox model contained 13 genera, including Acinetobacter, Brachybacterium, Exiguobacterium, Faecalibacterium, Finegoldia, Haemophilus, Kocuria, Lactococcus, Moraxella, Porphyromonas, Prevotella, Rhodococcus, Rothia. The genera with coef>0 in the models were considered risk factors, while those with coef<0 were considered protect factors. Kaplan-Meier survival curves were plotted to report the association between the survival probability and the abundance of specific genera. The strategy for grouping included dichotomization of abundance measured by CPM and the presence or not. The significance was examined by log-rank test and two stage hazard rate comparison. Combined with the clinical covariate such as sex, age, tumor stage and grade, the risk score was tested using univariate and multivariate cox to determine whether our risk score of microbial features could acted as an independent prognostic factor.

We attempted to determine the centrality among the genera involved in the cox model and to find the hub genera. The estimateNetwork function in the R package "bootnet"^29^ as used and the correlation between the features were visualized with the network plot. The influence of each genus was also measured by the indexes including “Strength”, “Closeness”, “Betweenness” and “ExpectedInfluence”.

### Mapping interaction between genera and host gene

We previously got the gene expression of 6 cohorts. To filter genes non-related to protein coding, we mapped the gene list to the human genome profile named ‘Homo_sapiens.GRCh38.109.chr.gtf.gz’ downloaded from the ENSEMBL website (http://asia.ensembl.org/index.html) and 19142 genes finally remained. The gene expression of TCGA was downloaded from the GDC portal (https://portal.gdc.cancer.gov/) and the data format was transformed to TPM.

To figure out the molecular change, especially the signaling pathway differentiation between the sub-group stratified by the risk score determined by selected microbial features in cox model, we performed the gene set enrichment analysis (GSEA). We used the GSEA function in the R package “clusterProfiler”^30^, and the KEGG, PID and REACTOME database were all included using R package “msigdbr”^31^. The p-values were corrected with the BH method.

We took the mantel test to characterize the correlation between interest genera and interest molecular pathways. The mantel_test function in the R package “linkET”^32^ (ref) was used. We dichotomized the genera into two clusters labeled as risk genera and protect genera. To score the immune related function, the single-sample gene set enrichment analysis (ssGSEA) method in the R package “GSVA”^33^ (ref) was used. The immune cell infiltration was assessed by the quantiseq method using deconvo_tme function in the R package “IOBR”^34^. As there were 15 patients who received the nivolumab immunotherapy and were recorded the objective response rate in the EGAD00001006029 (Cohort 2), we compared the differential genera between two groups, that were CB and NCB, using the Chi-Squared test. The prediction ability was adjudged by the area under curve (AUC).

To macroscopically evaluate the association between tumor microbiome composition and host gene expression, we performed Procrustes analysis. BC dissimilarity was calculated and then the nonmetric multidimensional scaling (NMDS) was used for dimension reduction. The reduced two dimensions or axes was input for the rotations and statistical testing in Procrustes analysis. Furthermore, we took the sparse CCA to identify group level correlations between paired host gene expression and microbiome data using the CCA function in the R package “PMA”^35^. The parameters were set as default. We processed the data before the analysis. The genus whose relative abundance was higher than 0.001 in at least 10% samples were kept, and the data was transformed to the centered log ratio (CLR) format for downstream analysis. We kept the genes whose expression was greater than 0 in half of the samples and then filtered out genes with low variance, using 25% quantile of variance across samples in each disease cohort as cut-off. These filtering resulted in a unique microbiome abundance matrix and host gene expression matrix per cohort for downstream analysis, including 12477 gene × 54 taxa in the EGAD00001000597 cohort, 11817 gene × 28 taxa in the EGAD00001006029 cohort, 12633 gene × 26 taxa in the GSE126964 cohort, 11406 gene × 26 taxa in the GSE151419 cohort, and 11492 gene × 60 taxa in the Renji cohort. As the number of tumor samples in GSE102101 was small, we didn’t performed sparse CCA in this cohort. After the sparse CCA, we got paired genus and genes clusters with significant correlation, and they were classified into a component. The genes in each component were implemented with pathway enrichment analysis. The significance was determined by Fisher’s exact test and BH method used for adjustment.

### Statistical analysis

All data analyses were conducted via RStudio software unless otherwise specified. Visualizations were performed using ggplot2 R package. Two group comparisons were done using Wilcoxon rank-sum test. Spearman’s correlations were calculated using cor.test function. The heatmap was created using Heatmap in “ComplexHeatmap” R package. In this paper, we used the following notation to indicate the significance levels of P-values: NS (P > 0.05), *0.05 < P < 0.01, **0.01 < P < 0.001, and *** P < 0.001.

### Transmission Electron Microscopy (EM)

A total of 20 ccRCC tissue blocks were subject to EM. Fresh tissues were carefully handled immediately after surgical removal. Blocks sliced 1mm³ in size were placed in a culture dish containing an electron microscope fixation solution. Samples were rinsed in 0.1M phosphate buffer (PB, pH 7.4). Samples were then placed at room temperature for 2 hours using 1% osmium tetroxide prepared in 0.1M phosphate buffer (PB, pH 7.4). Gradual dehydration was applied, and infiltration was conducted in a mixture of propylene oxide and Epon 812 resin (1:1) at 37°C overnight. Samples were inserted into an embedding mold filled with pure Epon 812 resin. The embedding mold underwent polymerization in a 60°C oven for 48 hours. Ultrathin sections (70nm) were cut from resin blocks using an ultramicrotome and placed on 200 mesh Formvar-coated copper grids. Copper grids with sections were stained in a 2% uranyl acetate-saturated alcoholic solution for 15 minutes. Following three rinses with ultrapure water, sections were stained with a lead citrate solution for 10 minutes. Copper grid sections were air-dried at room temperature overnight in a copper grid box. The grids were observed under a transmission electron microscope (HITACHI, HT7800).

### 16S rRNA staining

We performed 16S rRNA staining in 178 samples mounted on a tissue microarray (TMA) chip from the Huashan cohort with an established protocol reported by our group previously^19^. Briefly, thorough sterilization of hood, blades, and relevant instruments was carried out. Deparaffinized sections were dehydrated, and protease K was applied at room temperature. 100 μM of EUB338-cy5 probes (sequence: 5′– GCTGCCTCCCGTAGGAGT–3′) diluted in 1 μM working solution were applied and samples were finalized with DAPI (1:500) staining.

## RESULTS

### ccRCC has low-biomass and most ITB are contaminants

As most cohorts in this study were sequenced by bulk RNA-seq, we applied a tailored decontamination algorithm (**Fig 2A**). Analysis of these datasets revealed diverse bacteria present in ccRCC samples (**Fig S1**). Raw ITB reads took up ∼(1/2.00E-06) of total sequencing reads (**Fig S2A**) and showed a positive correlation with total reads (**Fig S2B**). 327 out of 545 genera survived after decontamination (**Fig S2C**). Our passes not only managed to filter out nonhuman reads (**Fig S2D**) but also showed an increase in proportion of common contaminants after decontamination, indicating that some bacteria, previously accepted as contaminants could be indwelling in ccRCC (**Fig S2E**). Relative abundance of non-human associated bacteria dropped consistently in all cohorts following our decontamination (**Fig S2F**). Common genera across cohorts after decontamination remained comparable either grouped by dataset or by sample (**Fig S3A-D**), whereas similar trend for bacterial read drop was noticed in cancer and normal tissue, respectively, further authenticating the remaining reads were true ITB in ccRCC (**Fig S3E-F**). Compositional atlas demonstrated by relative abundance, as expected, varied drastically across cohorts (**Figure S4A-D**). Despite so, two phyla, Proteobacteria and Firmicutes were present in all bulk-sequenced cohorts (**Fig S4**) and in scRNA-sequenced samples (**Fig S5, Table S1**). They were putatively present in diverse cells such as tumor cells (**Fig S5**). Furthermore, we then applied 16S rRNA-targeted FISH probe and EM imaging to 20 ccRCC tissue blocks, validating ITB existence in ccRCC (**Fig 2B-C**). We also attempted to culture 5 tissue blocks in aerobic and anaerobic conditions, but no bacterial growth was noted, supporting low biomass feature of ccRCC (data not shown). We then cross-referenced top-20 abundant genera in all cohorts and found 11 genera were present in ≥ 5 cohorts (**Fig 2D, Fig S5B, Table S2**). Interestingly, three genera including *Cutibacterium* were also present in TCGA cohort processed by both approaches (**Fig S5C**). Here, we concluded ccRCC harbored a low biomass of ITB and identified presence and composition of ITB which was possibly extracellular in ccRCC.

**Figure 2.**
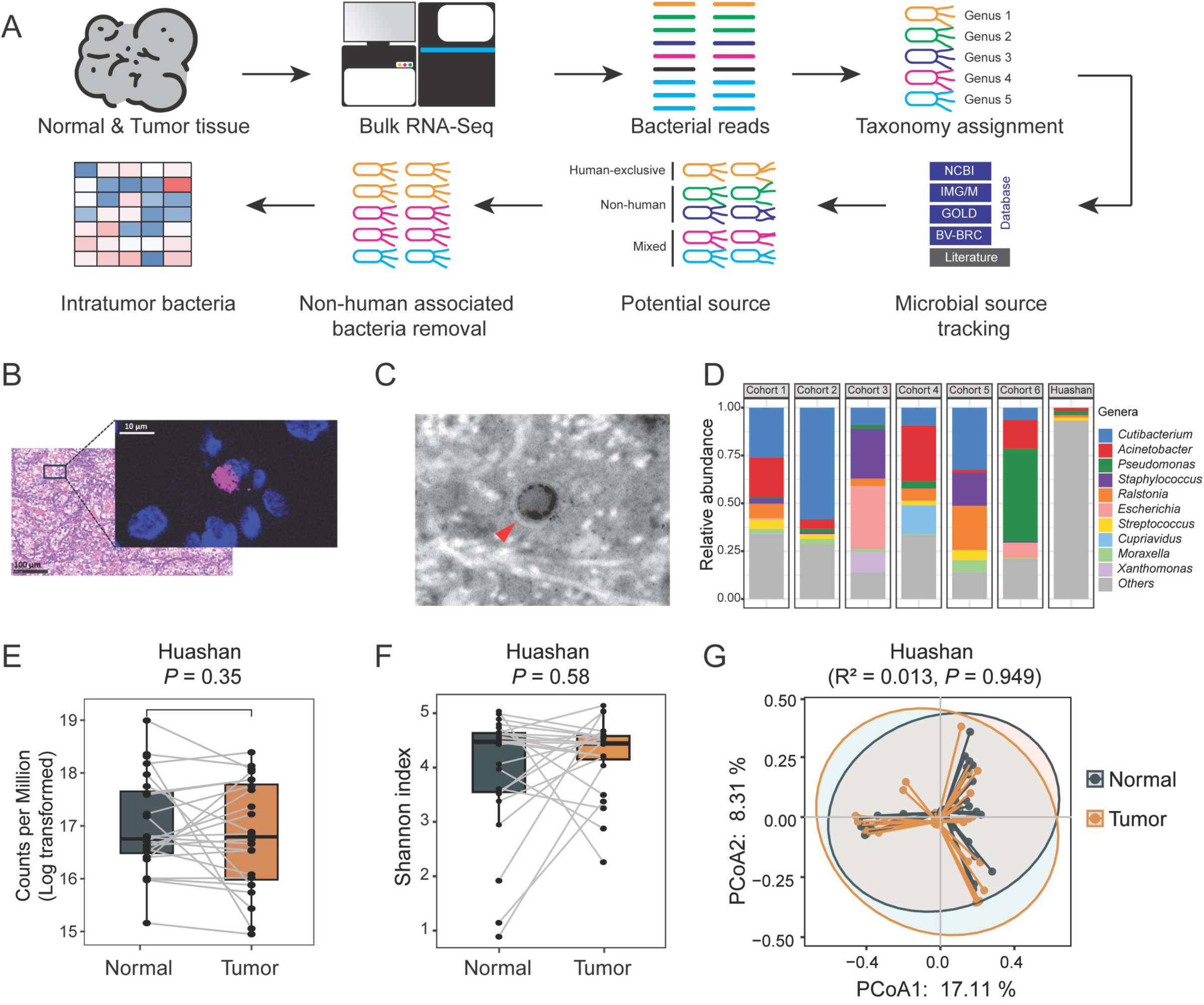
The presence of intratumor bacteria in ccRCC. (A) Flowchart illustrating the process of analyzing bulk RNA-seq data to identify intratumor bacteria. The analysis involves using bulk RNA-seq data from normal and tumor tissues. To track potential microbial sources, source annotations from bacterial genomes in databases such as NCBI, IMG/M, GOLD, and BV-BRC, as well as literature search, are retrieved. Bacteria associated with the human host are retained for constructing the intratumor bacteria matrix. (B) Representative images of fluorescence in situ hybridization (FISH) staining of 16S rRNA in tumor tissues of ccRCC. (C) Representative images of the presence of bacteria in the tumor tissues captured by transmission electron microscopy based on a total of 20 ccRCC tissue blocks. The red arrow indicates the object. (D) Stacked bar plot showing the proportion of genera present in at least five cohorts among the seven cohorts. Box plot showing the difference of (E) putative load (bacterial counts per million reads) and (F) Shannon index between 24 tumor and paired normal samples for comparison in Huashan cohort. The statistically significant difference was given by paired Wilcoxon rank-sum test. (G) Principal coordinate analysis (PCoA) for 24 paired tumor and normal samples in Huashan cohort, based on the Bray–Curtis dissimilarity. The *P* values were tested by Permutational multivariate analysis of variance (PERMANOVA).

### ITB does not differ between adjacent normal and cancer tissue in ccRCC

Using decontaminated reads (**Table S3**), we next probed clinical associations of ITB in three profiles: putative ITB load, ITB signature and individual ITB feature(s). Putative ITB load did not differ between paired normal and cancer tissue in all cohorts (**Fig 2E, Fig S6A**). Whereas cohorts that underwent RNA-seq could not be processed for alpha-diversity, we did not observe a difference in alpha-diversity in the Huashan cohort (**Fig 2F, Fig S6B-C**). Surprisingly, no differences in beta-diversity between normal and cancer tissue were observed in all cohorts (**Fig 2G, Fig S6D**). The only exception was TCGA_P cohort, which was challenged for its overinflated ITB reads (**Fig S6E**) and the alleged corrected version, TCGA_S cohort, again showed no difference (**Fig S6F**). We thus pursued whether individual ITB feature(s) was differentially distributed and was reproducible. Consistent with barren result of comparison between tumor and normal samples using Wilcoxon Test (**Fig S7A-C**), although the Random Forest identified 10 candidate differential ITB, this machine learning failed to validate those features with satisfactory predicting efficacy across the cohorts (**Fig S7D-E**). Again, the 10 features showed inconsistent trends in TCGA_P cohort and none was significantly different in TCGA_S cohort (**Fig S8A-B**). Here we show astonishingly that, contrary to most studies, differential ITB between adjacent normal and cancer tissue could very well be not present in ccRCC. Our findings highly suggested that most ITB in ccRCC could be inherent intra-tissue bacteria residing in kidney and only individual ITB features altered in abundance in cancer environment, supporting a passenger role of ITB in tumorigenesis stage of the disease.

### Putative ITB load and risk score predict survival in ccRCC

As expected, ccRCC could not be subtyped by ITB signature based on survival (**Fig S9A-B**). Indeed, ITB signature on the whole was not associated with any major clinicopathological parameters across cohorts (**Fig S10**). However, higher putative loads were associated with better overall survival (OS) in three cohorts available with OS profile (**Fig 3A**). In TCGA_S cohort that encompassed a small sample size, higher loads conferred a numerical better survival whereas TCGA_P cohort showed no difference, further questioning data processing in TCGA_P cohort (**Fig S11A**). Higher putative loads were solely associated with a better progression-free survival (PFS) in two cohorts, not reproducible in one of our original cohorts (cohort 6, Renji) and played a marginally protective role in TCGA_S cohort (**Fig S11B-C**). We then identified the compositional differences between patients with long and short survival, and the genera that coexisted and possessed consistent risk in univariate cox across three cohorts was used as input for LASSO and Cox model constructing (Fig S12A-B). The model identified a 7-genera ITB risk score predictive of OS in all three cohorts (**Fig 3B, Fig S12C-D, Table S4**) but not in either TCGA cohort (**Fig S12E, Table S5**). Specifically, *Actinomyces* and *Bifidobacterium* were protective ITB in ccRCC (**Fig S13**). Similar methodology was applied to PFS probing and a 13-genera risk score was generated (**Fig S14, Table S6**). Higher score predicted worsened PFS in all cohorts (**Fig 3C**) in which *Exiguobacterium* was a risk factor and *Rothia* was protective (**Fig S15**). Likewise, the results were not reproducible in either TCGA_P or TCGA_S cohort (**Fig S16, Table S7**). Whereas TCGA_P was problematic and TCGA_S consisted of only WGS samples, we here provided solid evidence that both ITB loads and features played a role in prognosis. This encouraged us to further investigate host interactions and treatment response.

**Figure 3.**
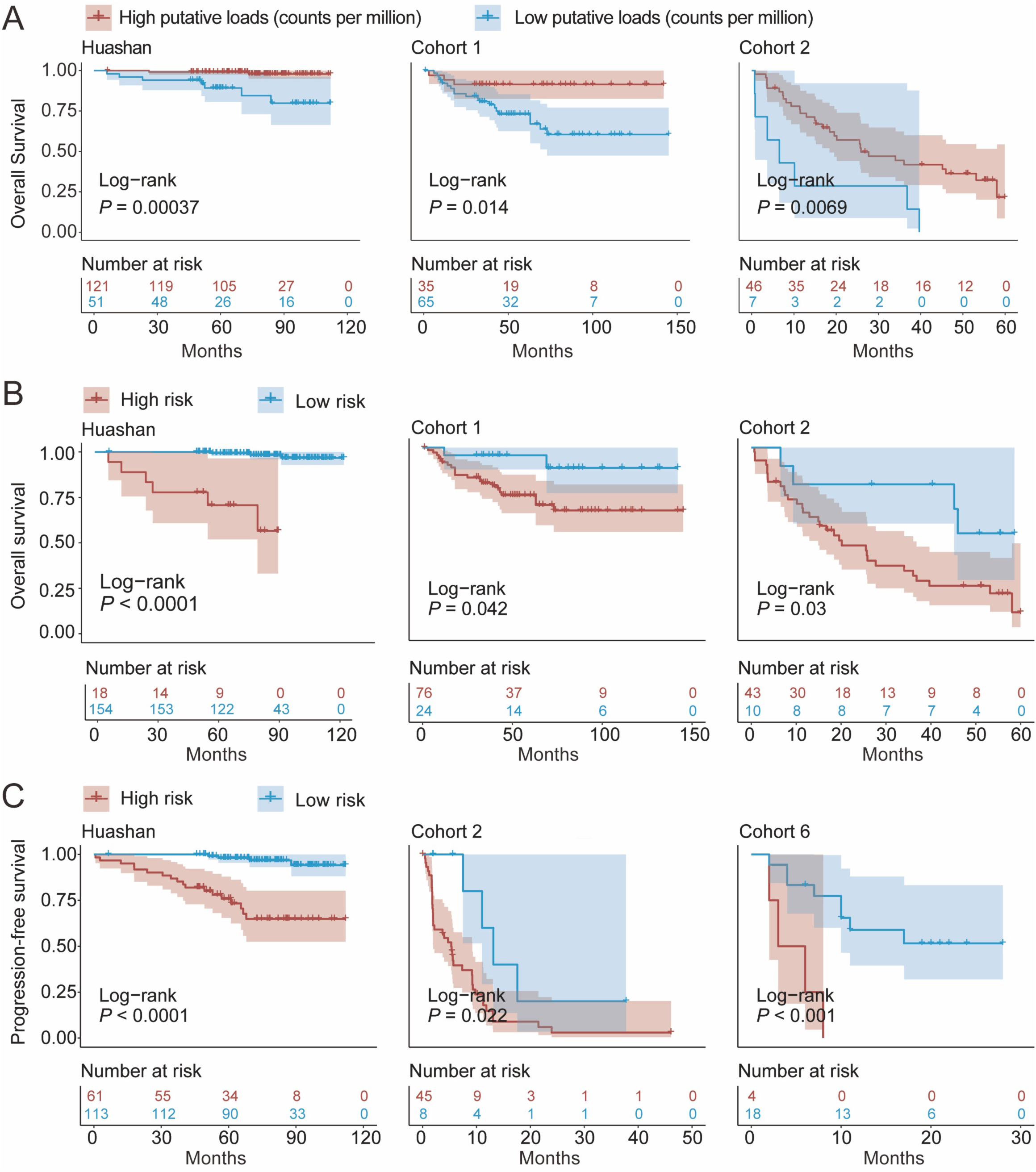
Putative ITB loads and risk score predict survival in ccRCC. Kaplan–Meier curves showing the overall survival probability for Huashan, Cohort 1, and Cohort 2 stratified by (A) putative loads and (B) risk score. (C) Kaplan–Meier curves showing the progression-free survival probability for Huashan, Cohort 2, and Cohort 6 stratified by risk score. *P* values were calculated using an unadjusted Log-Rank test.

### ITB is immune-related in ccRCC

Of exploratory interest, we investigated interactions between prognosis-related ITB (**Fig S12D, Fig S14C**) and found *Actinomyces* and *Rothia* being consistent hub ITB features across cohorts (**Fig S17**). When host interactions were incorporated, we found the immune response to be the sole consistently enriched program in ITB risk score-stratified patients across all cohorts (**Fig 4A, FigS18A-B, Table S8-11**). In reminiscence of inter-ITB interactions, ITB genera were associated with antigen presenting cell functions (co-inhibition and co-stimulation). (**Fig 4B, Fig S18C-D**). The risk score ITB showed in general negative correlation with pro-cancer immune infiltrates (**Fig 4C**). Specifically, absence of protective ITB *Actinomyces*, *Rothia* and *Bifidobacterium* were associated with M2 polarization of macrophages (**Fig 4D, Fig S18E-G**). Nonetheless, those three features were not associated with response to immune checkpoint inhibition and we identified *Anaerococcus* and *Corynebacterium* enriched in ccRCC with complete response (CR) to Nivolumab (**Fig 4E-F**). Lastly, we profiled host interaction using Sparce CCA and three out of five cohorts showed significant host gene-ITB interaction (**Fig S19A**). Besides immune, we also noted Ribosome signaling was associated with some microbiota across all cohorts (**Fig S19B-G**). Here, we showed ITB was associated with host immune response in particular protective ITB that were associated with decreased immune escape.

**Figure 4.**
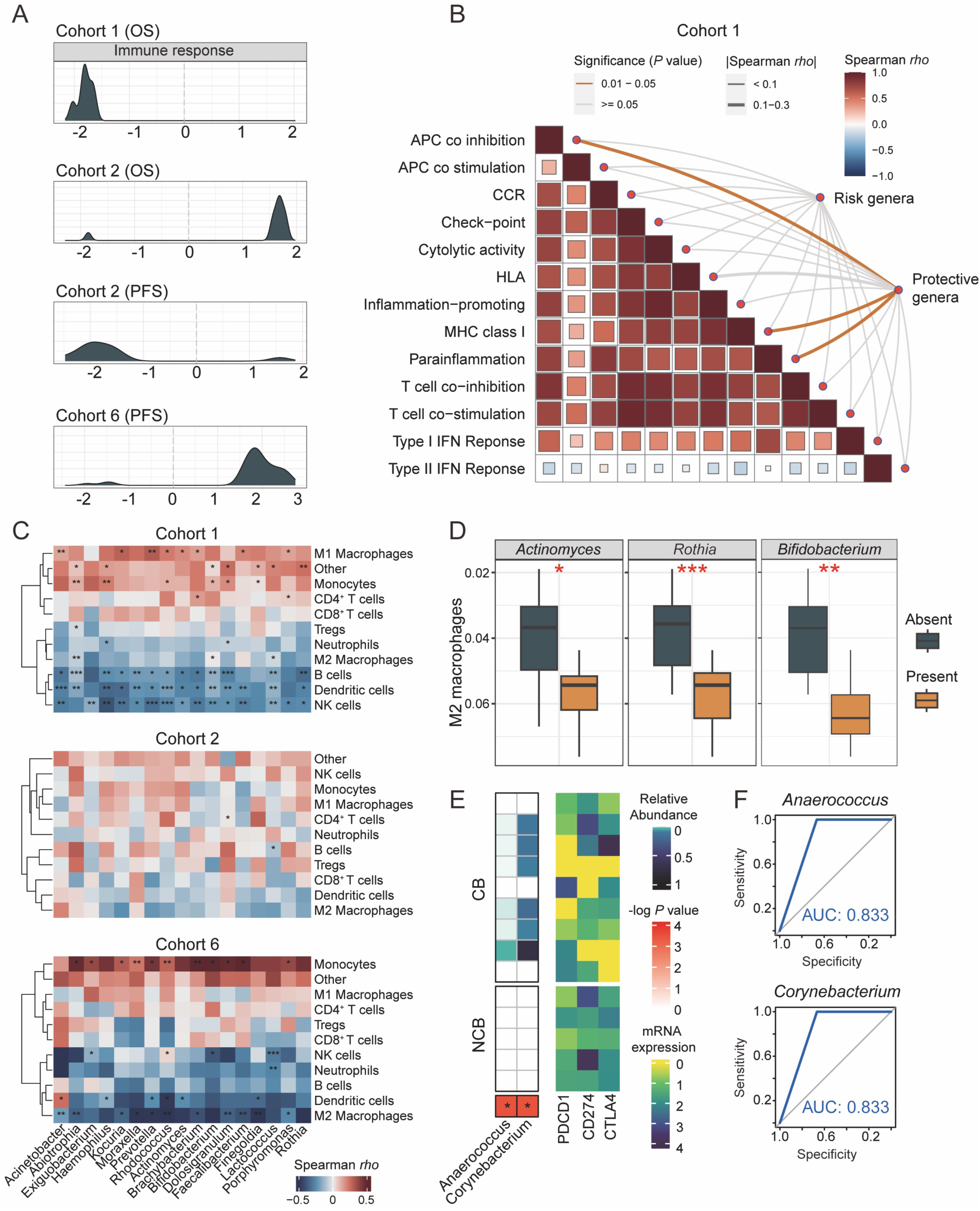
ITB is immune-related in ccRCC. (A) The density curves represent the distribution of the immune-related pathways that were significantly enriched between the two stratified groups using gene set enrichment analysis. The horizontal axis indicated the NES of the GSEA result. The stratification was the same as the previous result, that is the overall survival-related risk group in Cohort 1 and 2, and progression-free survival-related risk group in Cohort 2 and 6 from top to bottom. (B) The result of the Mantel test showing the interaction between genera community and potential immune function and the Spearman method was used. The thickness of the curve indicated the absolute value of the spearman rho, and the significant connection was yellow colored. Each block represented the correlation among the immune functions, and a redder color meant a greater rho. (C) Heatmap showing the correlation between specific genus and infiltration scores of immune cells in Cohort 1, Cohort 2, and Cohort 6. (D) Box plot exhibiting the level of M2 macrophage polarization in the presence or absence of *Actinomyces*, *Rothia*, *Bifidobacterium* from left to right in Cohort 6. The Wilcoxon Test was used for comparing the relative abundance between tumor and normal. (E) The heatmap at left showed the relative abundance of the differential genera in abundance in the patients with clinical benefit (CB) and non-clinical benefit (NCB) using Chi-Squared Test. The heatmap at the right indicated the mRNA expression of genes *PDCD1*, *CD274*, and *CTLA4*. (F) The ability of the abundance of *Anaerococcus* and *Corynebacterium* to predict the clinical benefit was visualized by the receiver operating characteristic curve and measured using the area under the curve (AUC).

## DISCUSSION

Our study encompassed thus far the largest number of ccRCC cases subject to ITB detection. In comparison to previous smaller studies^14–17^, several ITB features appeared to be ubiquitously present at high relative abundance including Proteobacteria and Firmicutes at phylum level and *Pseudomonas*, *Acinetobacter* and *Staphylococcus* at genus level. Lack of difference in ITB loads, composition or diversity between normal and cancer tissue was one of our major findings. Though it was previously reported by Wang et al, we initially considered it to be a result of lack of any decontamination in their study^16^. Given that ITB features associated with prognosis were not amongst the top abundant ones, we speculate that ITB could be sporadic and commensal, not just in ccRCC but also in kidney.

Though our 7-genera panel appeared to perform consistently in all cohorts, we are yet to conclude a pathogenic mechanism regarding a single ITB. Like in genetic association studies, prognostic panel composed of multiple genes serves as a biomarker simply because none of the individual gene is statistically powerful enough to generate a reproducible survival difference and any attribution of a single element should be supported by mechanistic analyses by cell or animal modeling. Likewise, our ITB panel solely represents the prognostication of the microbial community. Moreover, our ITB panel was only aggregated at genus rather than species level, further against overinterpretation.

The causation between ITB and renal tumorigenesis remains unknown^36^. Whether those prognostic ITB are still commensal or, playing driving roles alongside tumor progression depends on human microbiota-associated murine models (HMAMMs) and microbe-phenotype triangulation (MPT)^36^. Unfortunately, there are currently no transgenic murine models for ccRCC^37^ and culturomics from animal models is therefore inapplicable^38^. It was surprising that most prognosis-associated ITB features were protective and so were high putative loads, contrary to many oncobiome studies. We did not evaluate absolute ITB loads in our own cohorts as loading could not be accurately calculated in transcriptome datasets. However, given that recent study points out that absolute, rather than relative abundance plays more important role in microbiome study^39^, and that load is prognostic in nasopharyngeal cancer (NPC)^40^, we are now setting up a new line to evaluate association between absolute ITB loads and prognosis. We did not put much effort into imaging ITB. For low-biomass cancer, both LPS and FISH staining could harbor magnified signals from extra-tumor bacterial contamination^41^. We consider multiple sequencing platforms together with FISH signaling adequate to prove the existence of ITB. Of note, we did not identify any intracellular bacteria either by EM or scRNA-seq. This could either be inherent biology of ccRCC or be a result of extreme low biomass of kidney as we successfully identified ITB in all 10 samples of bladder cancer undergoing scRNA-seq in another companion project (data not shown).

Recent debate over the landmark cancer microbiome study by Poore et al^1^ has drawn much attention in the oncobiome community. In their recent report^25^, Salzberg’s team reasoned two major points that Poore’s data should be interpreted with caution, including contamination of human reads into microbial signaling and overinflation of microbial reads by machine learning. We owe great thanks to the Salzberg team for providing us KIRC WGS data processed with their protocol for reproduction and validation of our own findings. Even with the very limited sample size, our model showed a numerical OS prediction. The reason Salzberg’s team did not process RNA-seq samples was that they considered poly(A)-based transcriptomes could not capture microbial signals. However, half of our cohorts were poly(A)-based transcriptomic datasets and we were able to retrieve effective reads therein. In fact, most ITB studies using scRNA-seq were also able to capture effective reads given the very few cells compared with bulk sequencing. The “poly(A)” problem in the intratumor microbiome has also been thoroughly discussed^42,43^ and our findings undoubtedly further supported the notion.

Last but importantly, we show that certain ITB feature is associated with cancer immunity and response to Nivolumab in ccRCC, in reminiscence of recent trial modulation gut microbiome in metastatic ccRCC patients receiving Nivolumab plus ipilimumab therapy^44^. The protective ITB in our findings are closely related to decreased immune escape, e.g., inhibition of antigen presentation and decreased M2 polarization, both showing pro-inflammatory effects. Interestingly, ITB with different clinical associations seldom overlap and we have not identified such an “omnipotent” ITB in ccRCC. Despite so, *Corynebacterium* is of interest as its abundance ranks top 20 in most cohorts and is associated with Nivolumab response. *Bifidobacterium* supplement has been shown in trial that augments ICI response in metastatic ccRCC patients and our findings that intratumor *Bifidobacterium* was protective shed light on the thus far elusive mechanism of this gut-tumor asix^44^. We did not analyze ITB in ccRCC patients treated with angiogenesis-targeting therapy though there are a handful of datasets available, as angiogenesis was not amongst the MAMPs we identified (**Fig S19B**). Given that combination therapy has become the mainstay of metastatic ccRCC treatment, we are now setting up an ITB analysis in such samples.

## CONCLUSION

ITB exists in ccRCC. High ITB loads predicted better survival. We also developed a robust ITB score predictive of prognosis regardless of sequencing tech, sample processing or racial disparity. Those parameters and panels serve as novel biomarkers for ccRCC.

## DECLARATION

### Ethics approval and consent to participate

Informed consent was obtained for all patients and the study was approved by Huashan Institutional Review Board (HIRB2011-009; HIRB2023-908) and Renji Hospital, School of Medicine, Shanghai Jiao Tong University (KY2023-049-B).

### Data availability statement

Read counts of un-decontaminated ITB have been deposited at China National Center for Bioinformation (GSA: CRA011414) that are publicly accessible at https://ngdc.cncb.ac.cn/gsa. Request for TCGA-KIRC data processed by Salzberg et al should be addressed to Prof. Steven Salzberg.

### Authors’ contributions

Conceptualization: CF, WZ, YxL, HJ; Methodology: CF, LT, YL, DZ, YxL; Validation: LT, YL, DZ, HJ; Investigation: CF, LT, YL, DZ, RZ, YS, TG; Original Draft: CF, YL, YC

## Acknowledgments

This study was sponsored in part by the National Natural Science Foundation of China (Grant No. 81874123 and 82273248). We owe great thanks to the Salzberg team for sharing their data and grant for our use for publication. This study makes use of data generated by the Department of Pathology and Tumor Biology, Kyoto University, and Dr. Seishi Ogawa was highly appreciated. We acknowledge Bristol-Myers Squibb Company (BMS) as the source of the EGAD00001006029 cohort data.

