## Supplemental Figures for "Intratumor bacteria is associated with prognosis in clear-cell renal cell carcinoma"

**Supplementary Figures**

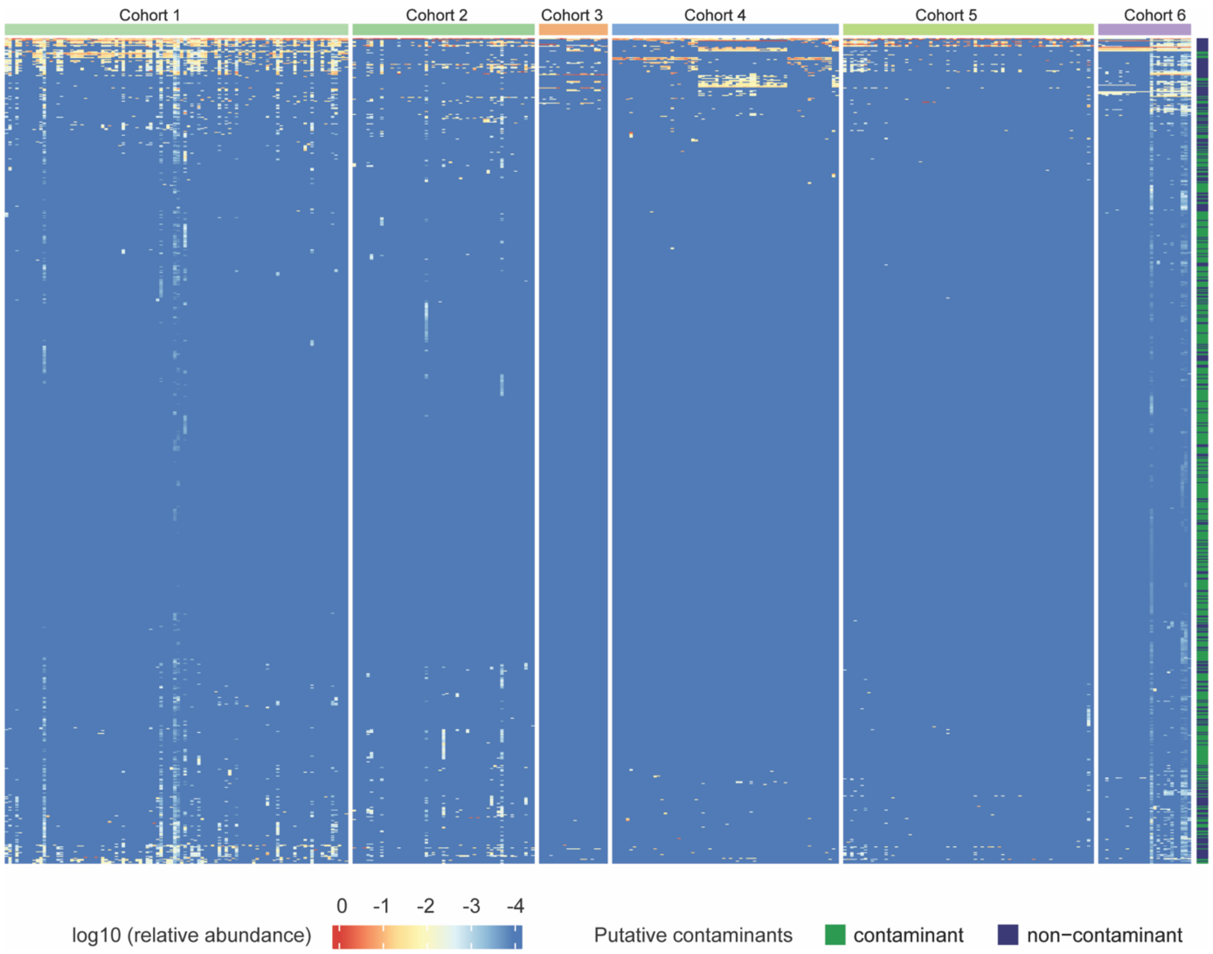

**Figure S1. Bacterial genera detected in bulk RNA-seq data from six cohorts**

The analysis of bulk RNA-seq data from six cohorts detected a total of 545 bacterial genera. Of these, 327 were identified as non-contaminant genera, while the remaining 218 were categorized as contaminant genera.

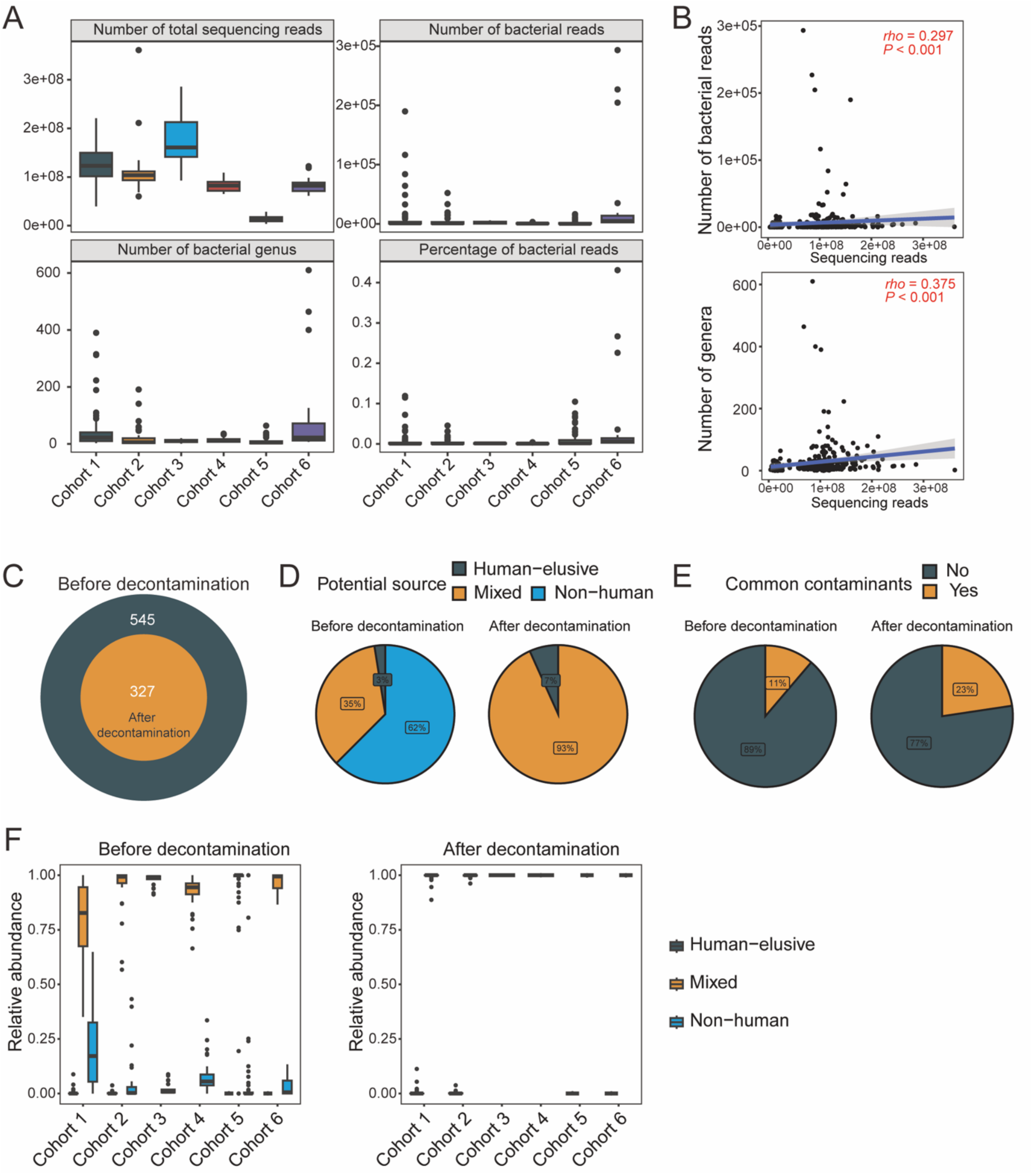

**Figure S2. Decontamination statistics for bulk RNA-seq data from six cohorts**

(A) Box plot displaying the distribution of the number of sequencing reads, bacterial reads, bacterial genera, and percentage of bacterial reads in RNA-seq data from six cohorts before decontamination. (B) The Spearman correlation between total sequencing reads and detected bacterial reads, as well as total sequencing reads and the number of detected genera. The blue line represents the fitted linear regression line, with a grey shading representing the 95% confidence interval. (C) A Venn diagram illustrating the number of detected bacterial genera before and after decontamination. (D) Pie charts showing the proportion of overall bacterial genera belonging to different potential sources before (left) and after (right) decontamination, as well as the proportion of remaining genera belonging to common contaminants (E) before (left) and after (right) decontamination. (F) Box plots showing the distribution of the total relative abundance of genera of different potential sources in six cohorts with RNA-seq data, before (left) and after decontamination (right).

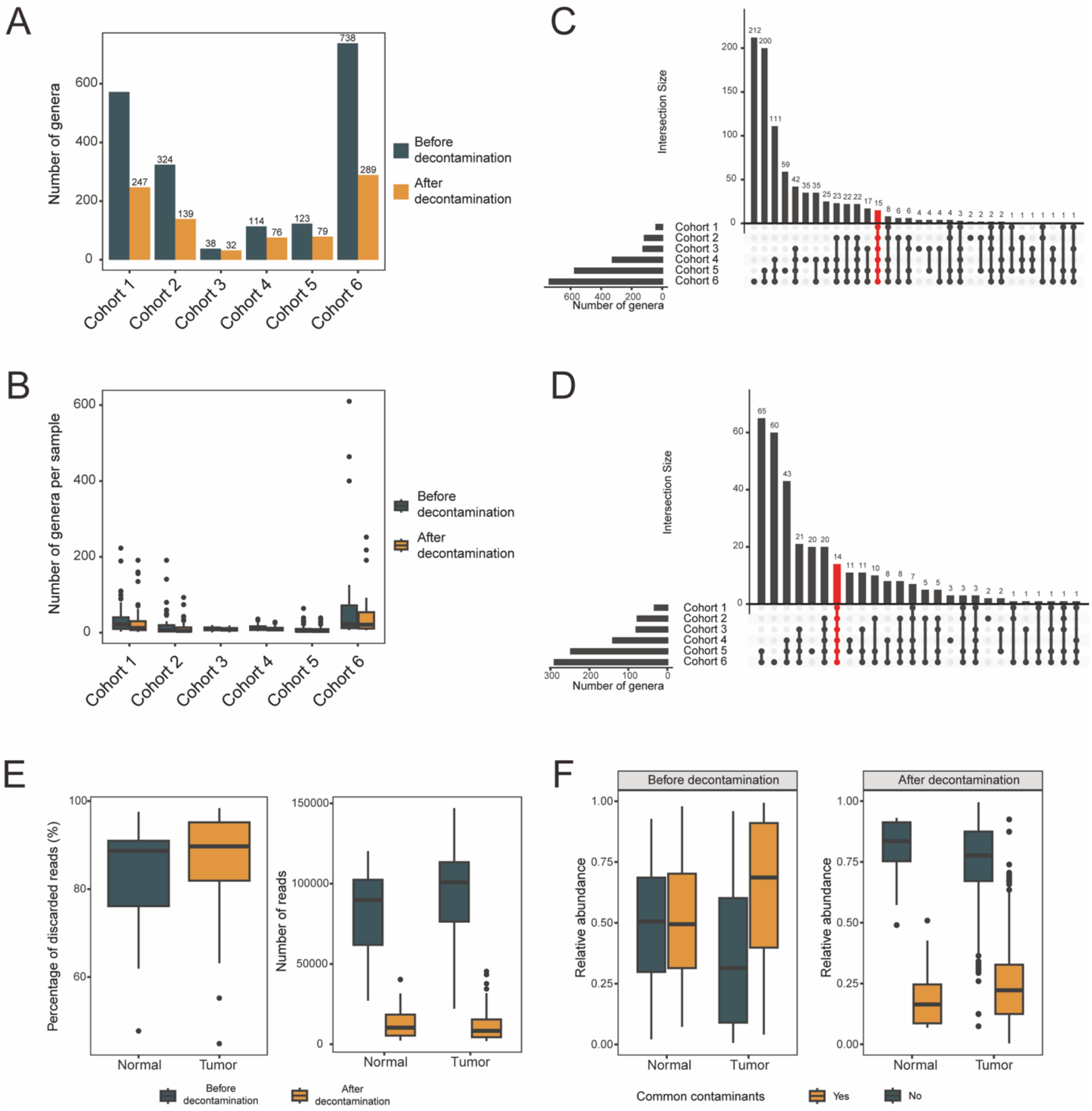

**Figure S3. Decontamination statistics for bulk RNA-seq data and 16S rRNA sequencing data**

(A) Bar plots showing the total number of detected genera in six RNA-seq cohorts before and after decontamination. (B) Box plots depicting the distribution of detected genera in six RNA-seq cohorts before and after decontamination. UpSetR plots illustrating the intersection of detected genera in six RNA-seq cohorts before (C) and after (D) decontamination. (E) Box plots showing the distribution of the percentage of discarded reads after decontamination (left), as well as the number of remaining bacterial reads before and after decontamination (right) in 16S rRNA sequencing data from Huashan cohort. (F) Box plots displaying the relative abundance of genera belonging to common contaminants before (left) and after decontamination (right) in 16S rRNA sequencing data from Huashan cohort.

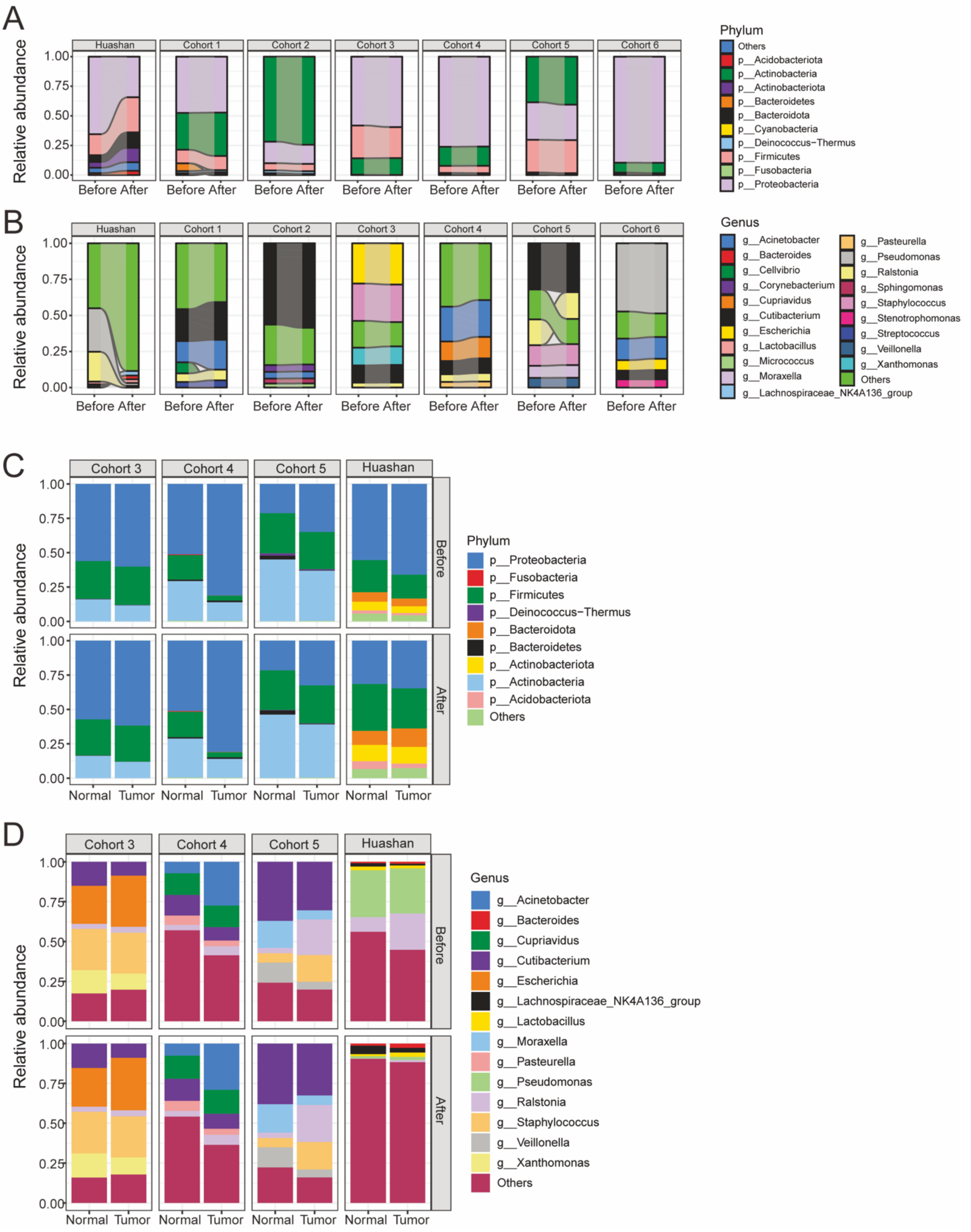

**Figure S4. Relative abundance of intratumor microbiota in ccRCC**

Alluvial plots of the phylum (A) and genus (B) level relative abundance of bacteria communities in seven datasets. Stacked bar plot of the phylum (C) and genus (D) level relative abundance of bacteria communities, with being stratified by normal and tumor tissues. Only the top five abundant phyla/genera in each dataset are shown. Before, before decontamination; After, after decontamination.

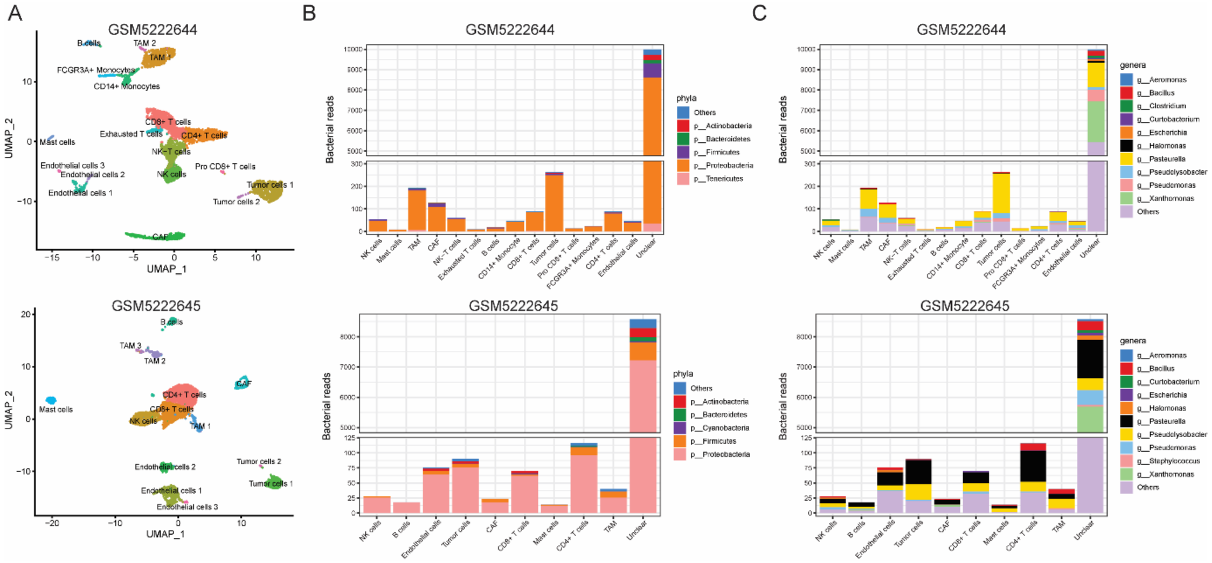

**Figure S5. Single-cell RNA sequencing data analysis**

(A) Uniform manifold approximation and projection (UMAP) plot showing cell origins by color. Stacked barplot illustrating the bacterial reads of bacterial phyla (B) or bacterial genera (C) detected in different cell types. Cells that cannot be assigned to a specific cell type due to low quality are grouped into “Unclear”.

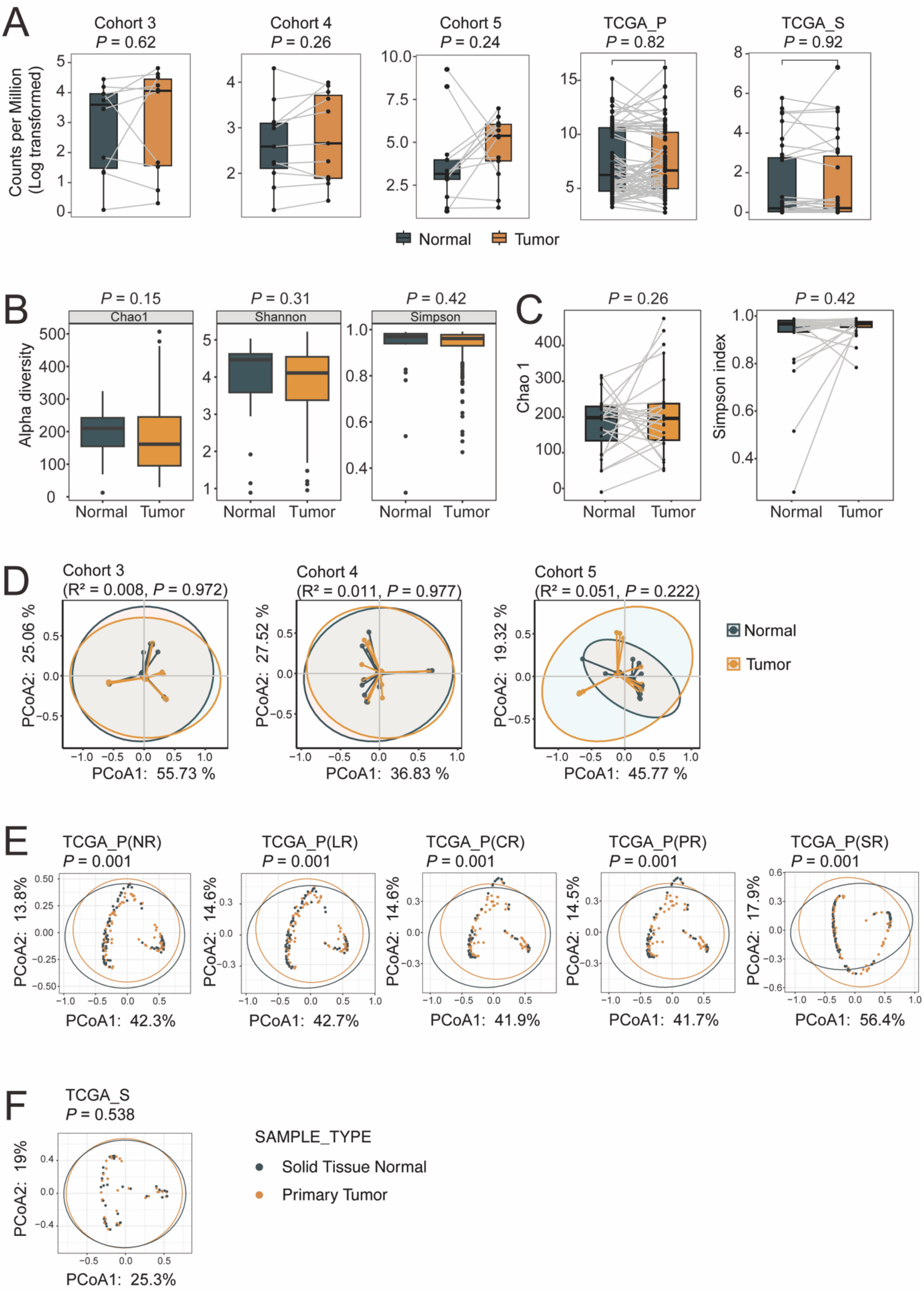

**Figure S6. Diversity comparison between tumor and normal samples in ccRCC**

(A) Box plot showing the difference of putative load (bacterial counts per million reads) between paired tumor and normal samples in Cohort 3, Cohort 4, Cohort 5, TCGA_P, and

TCGA_S. (B) Alpha diversity of all normal and tumor samples in Huashan dataset (16S rRNA sequencing data). Statistical significance was given by two-tailed Wilcoxon rank-sum test. (C) Alpha diversity of paired normal and tumor samples in Huashan dataset (16S rRNA sequencing data). Statistical significance was given by paired Wilcoxon rank-sum test. (D) Principal coordinate analysis (PCoA) for paired tumor and normal samples in Cohort 3, Cohort 4, and Cohort 5 and (E) TCGA_NR, TCGA_LR, TCGA_CR, TCGA_PR, TCGA_SR and (F) TCGA_S based on the Bray–Curtis dissimilarity. The P values were tested by Permutational multivariate analysis of variance (PERMANOVA).

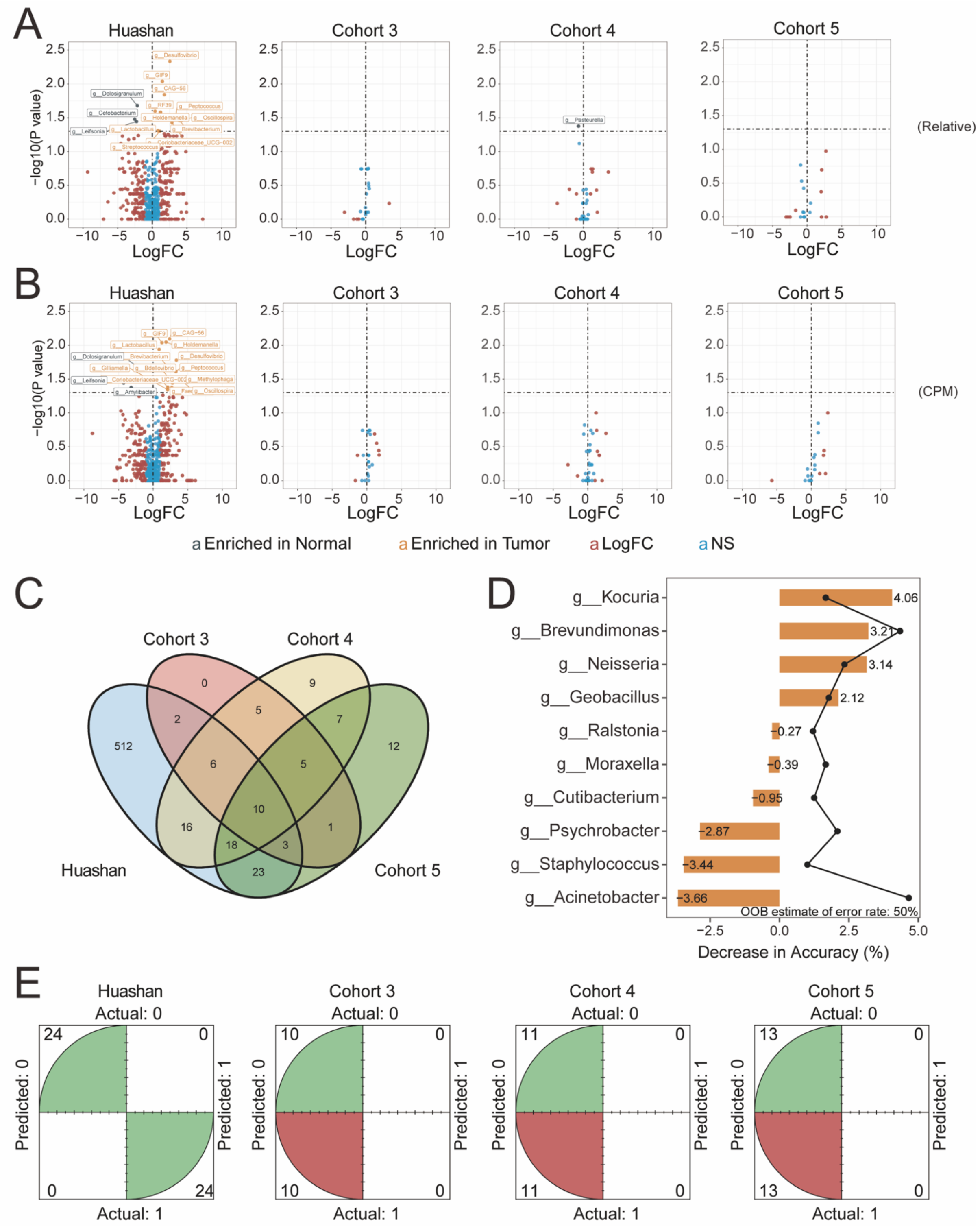

**Figure S7. Identifying genera differentially present in tumor and normal samples**

Volcano plot showing the genera with significantly differential abundance represented by (A) relative abundance and (B) counts per million (CPM) between tumor and paired normal samples of ccRCC in Huashan, Cohort 3, Cohort 4, Cohort 5 using Wilcoxon Test. The labels “Enriched in Normal” and “Enriched in Tumor” represent the genera that were significantly enriched in normal and tumor samples, respectively. The label “LogFC” indicates |LogFC| > 1 and *P* value < 0.05. The label “NS” indicates no statistical significance. (C) Venn plot showing the intersection of the genera among Huashan, Cohort 3, Cohort 4, Cohort 5. (D) The importance of feature genera selected by Random Forest for identifying key factors to distinguish the tumor samples from the normal. Huashan cohort was used for training. The horizontal axis represented the accuracy, and the black point indicated the Gini index. A feature with higher decrease in accuracy and higher Gini index was considered more important. (E) Four-quadrant diagram showed the predicting ability of the random forest model. The green indicated correct while the red was wrong.

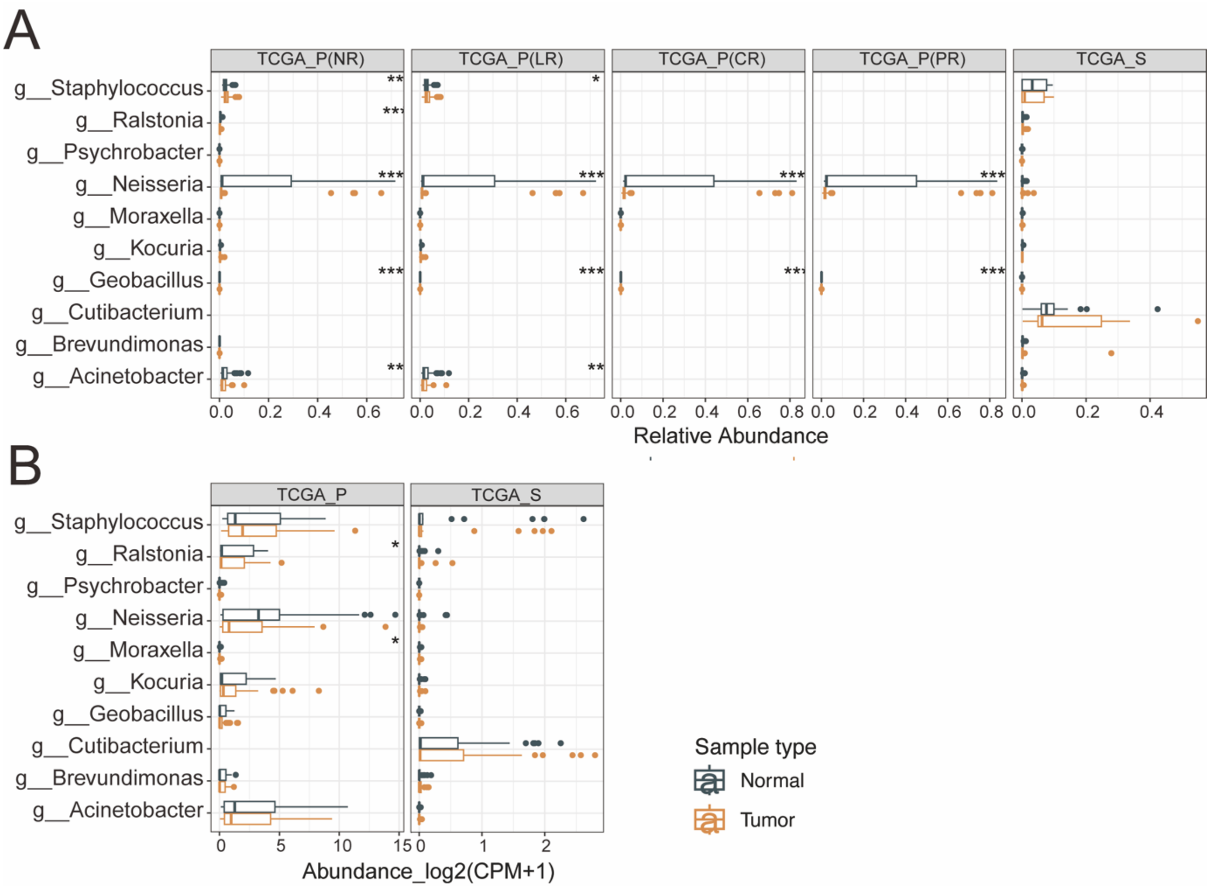

**Figure S8. Abundance of ten selected features in TCGA**

The bar plot showing the (A) relative abundance and (B) counts per million (CPM) of the selected ten feature genera in TCGA_P and TCGA_S. As no significantly differential genus was detected in TCGA_P (SR), the data for that is not shown. The difference between tumor and normal samples was compared using the Wilcoxon Test.

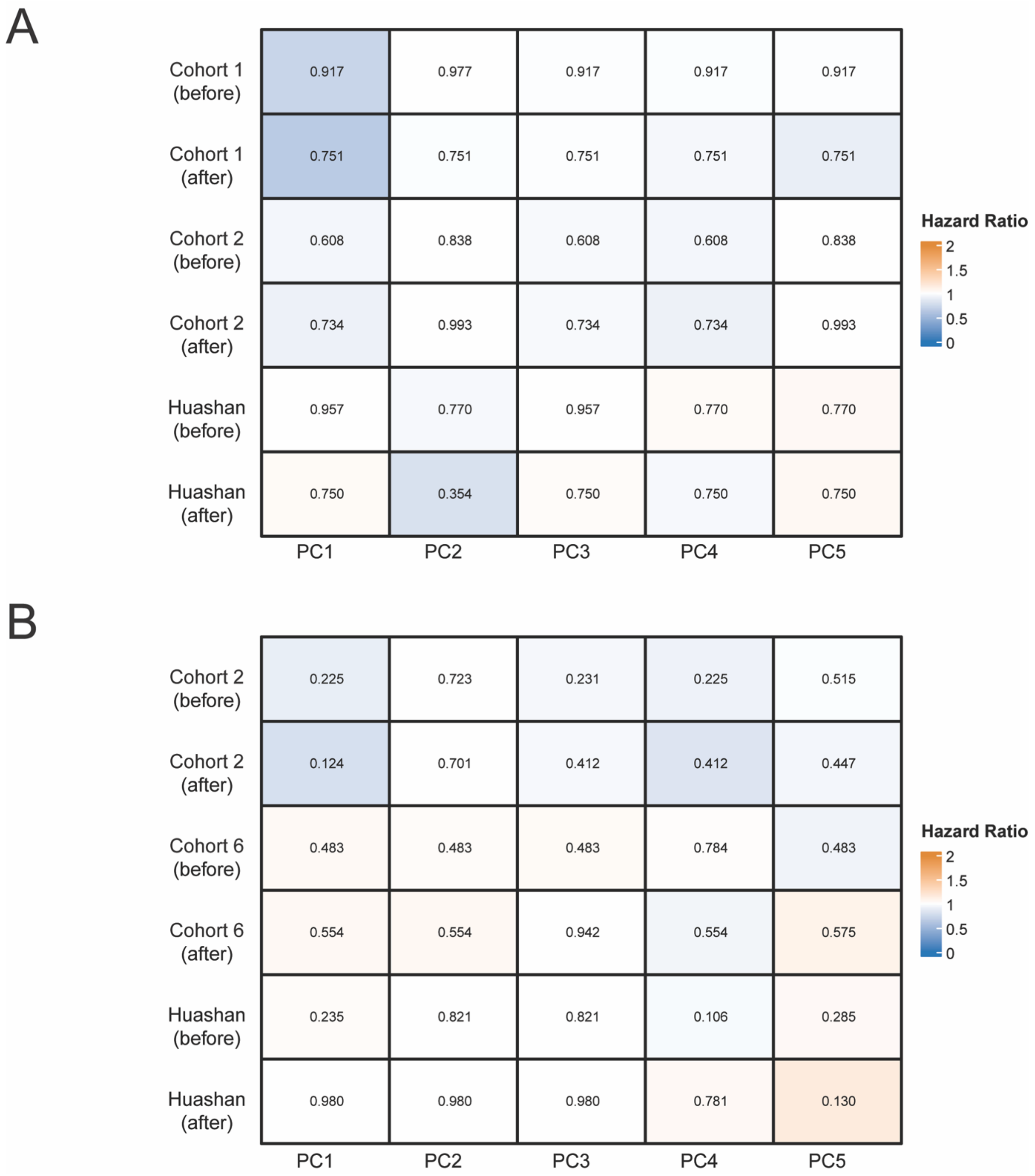

**Figure S9. Association of microbiome profiles with overall survival and progression-free survival**

Heatmaps showing the associations between the first five principal components (PCs) of intratumor bacteria profile and overall survival (A) or progression-free survival (B), which were examined by Cox proportional hazard regression models. The values in heatmaps represent *P* values. Before, before decontamination; after, after decontamination.

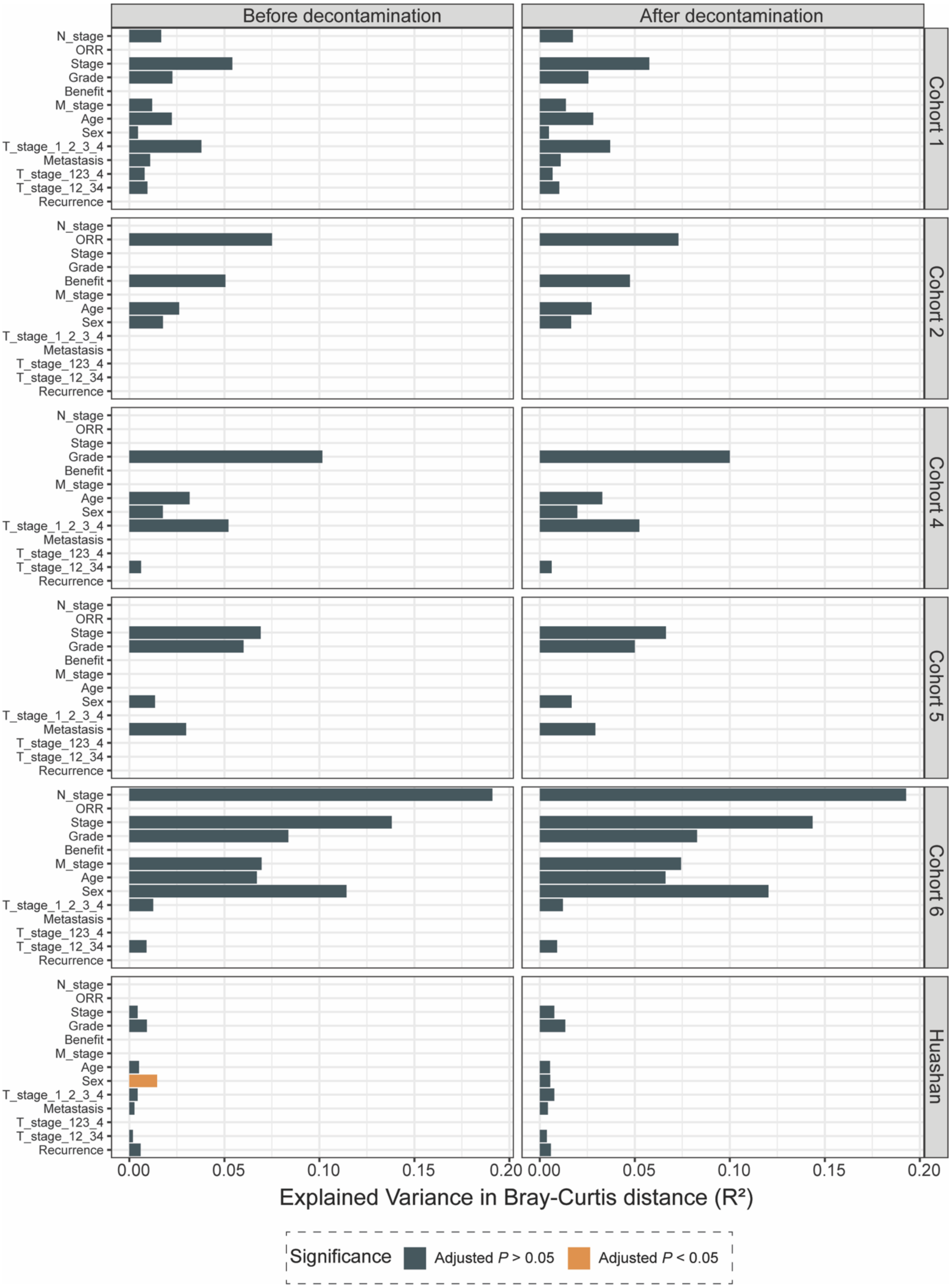

**Figure S10. Factors associated with intratumor bacteria composition in tumor tissues**

The bar plot indicates the explained variation of each factor in the interindividual variation of intratumor bacteria composition. PERMANOVA analysis with 999 permutations, based on Bray-Curtis dissimilarity, was performed to examine the effects of clinical factors on microbial communities. All *P* values were further adjusted for multiple comparisons with the FDR (false discovery rate) method. The clinical factors with FDR-adjusted *P* value < 0.05 were considered confounding factors. T_stage_1_2_3_4 represents patients being grouped into four groups (T stage1, 2, 3, 4); T_stage_123_4 represents patients being grouped into two groups (T stage1, 2, 3 vs T stage 4); T_stage_12_34 represents patients being grouped into two groups (T stage1, 2 vs T stage 3, 4).

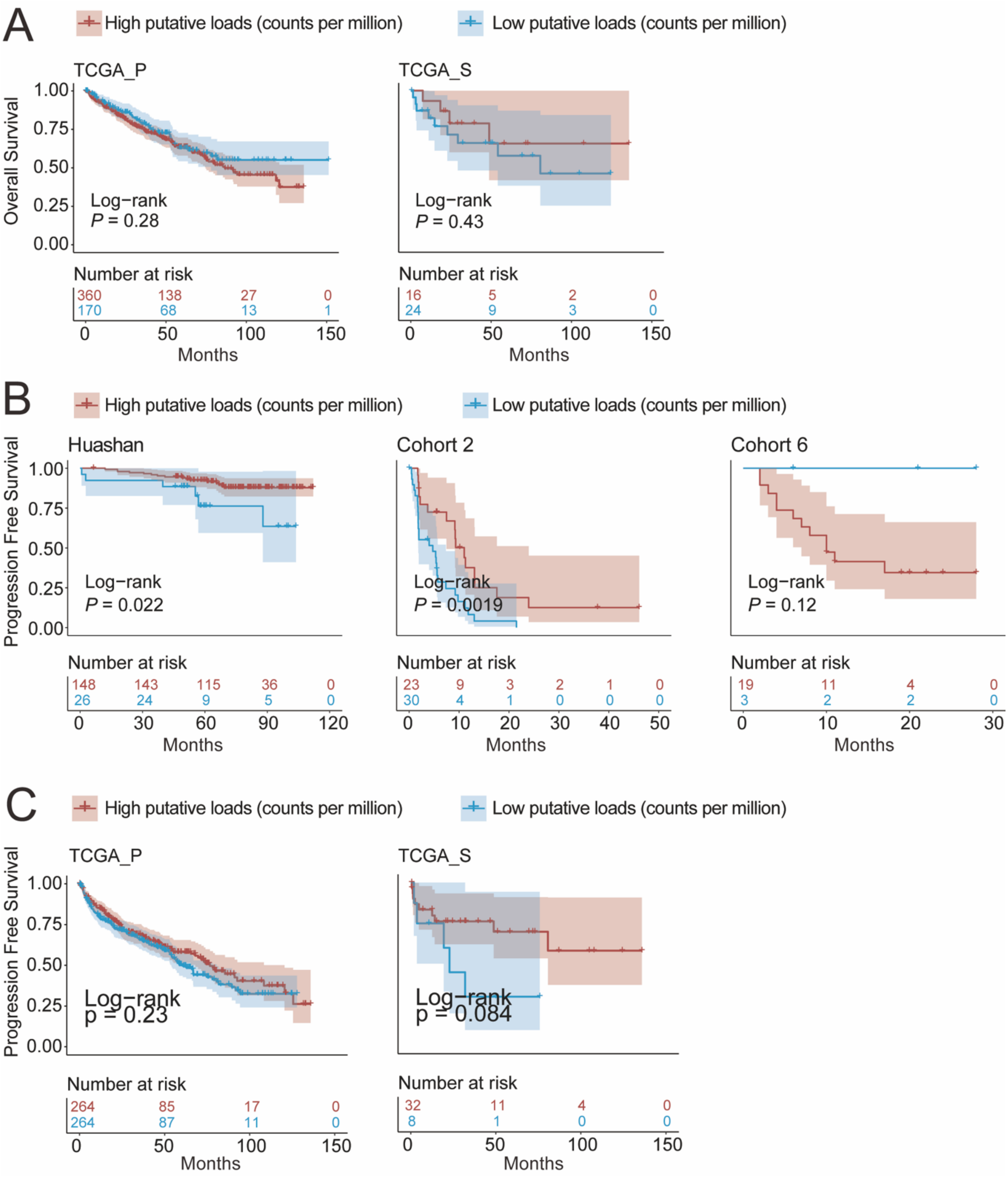

**Figure S11. Putative ITB load is associated with prognosis in ccRCC**

(A) Kaplan–Meier curves showing the overall survival probability for TCGA_P and TCGA_S stratified by the putative intratumor bacterial load. Kaplan–Meier curves showing the progression-free survival probability for (B) Huashan, Cohort 2, Cohort 6 (C) TCGA_P and TCGA_S stratified by putative intratumor bacterial load. *P* values were calculated using unadjusted Log-Rank test.

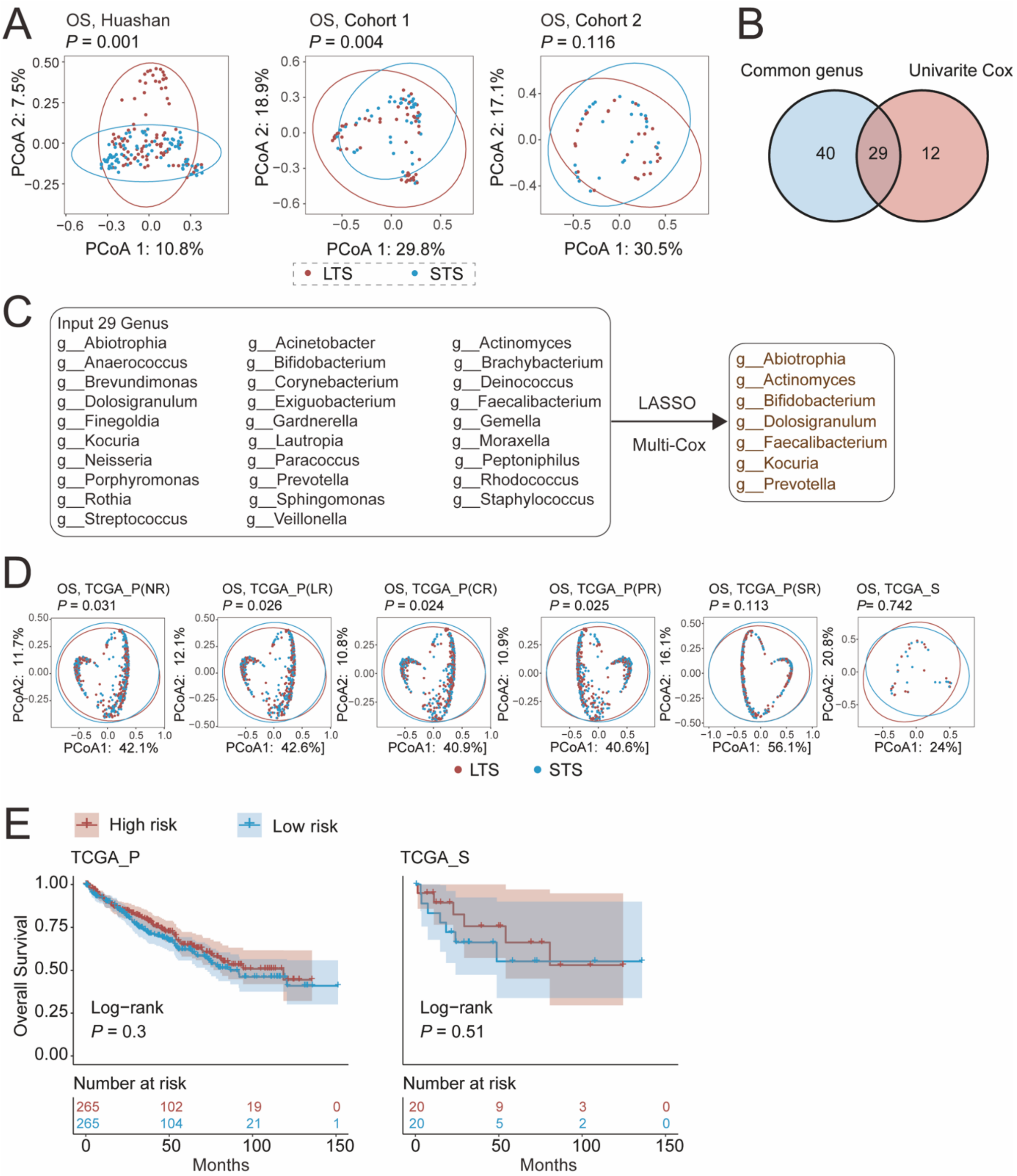

**Figure S12. Construction of microbial risk score for overall survival in ccRCC**

(A) Principal coordinate analysis (PCoA) for patients with long-term survival (LTS) and short-term survival (STS) of OS in Huashan, Cohort 1 and Cohort 2, based on the Bray–Curtis dissimilarity. The *P* values were tested by Permutational multivariate analysis of variance (PERMANOVA). (B) Venn plot showing the intersections between common genera and the genera with consistent HR in univariate cox for OS in the Huashan, Cohort 1 and Cohort 2. (C) The diagram presenting the input 29 genera for the first step: LASSO. Then the selected features by LASSO were delivered to construct a Cox model. Finally, we got a microbial community used for calculating a risk score for OS and each feature contributed positively or negatively to the score measured by the index "coef". The genera with coef > 0 were classified as risk genera and those with coef < 0 were classified as protective genera. (D) Principal coordinate analysis (PCoA) for patients with long-term survival (LTS) and short-term survival (STS) of OS in TCGA_P(NR), TCGA_P(LR), TCGA_P(CR), TCGA_P(PR), TCGA_P(SR), and TCGA_S, based on the Bray–Curtis dissimilarity. (E) Kaplan–Meier curves showing the overall survival probability for TCGA_P and TCGA_S stratified by risk score. *P* values were calculated using unadjusted Log-Rank test.

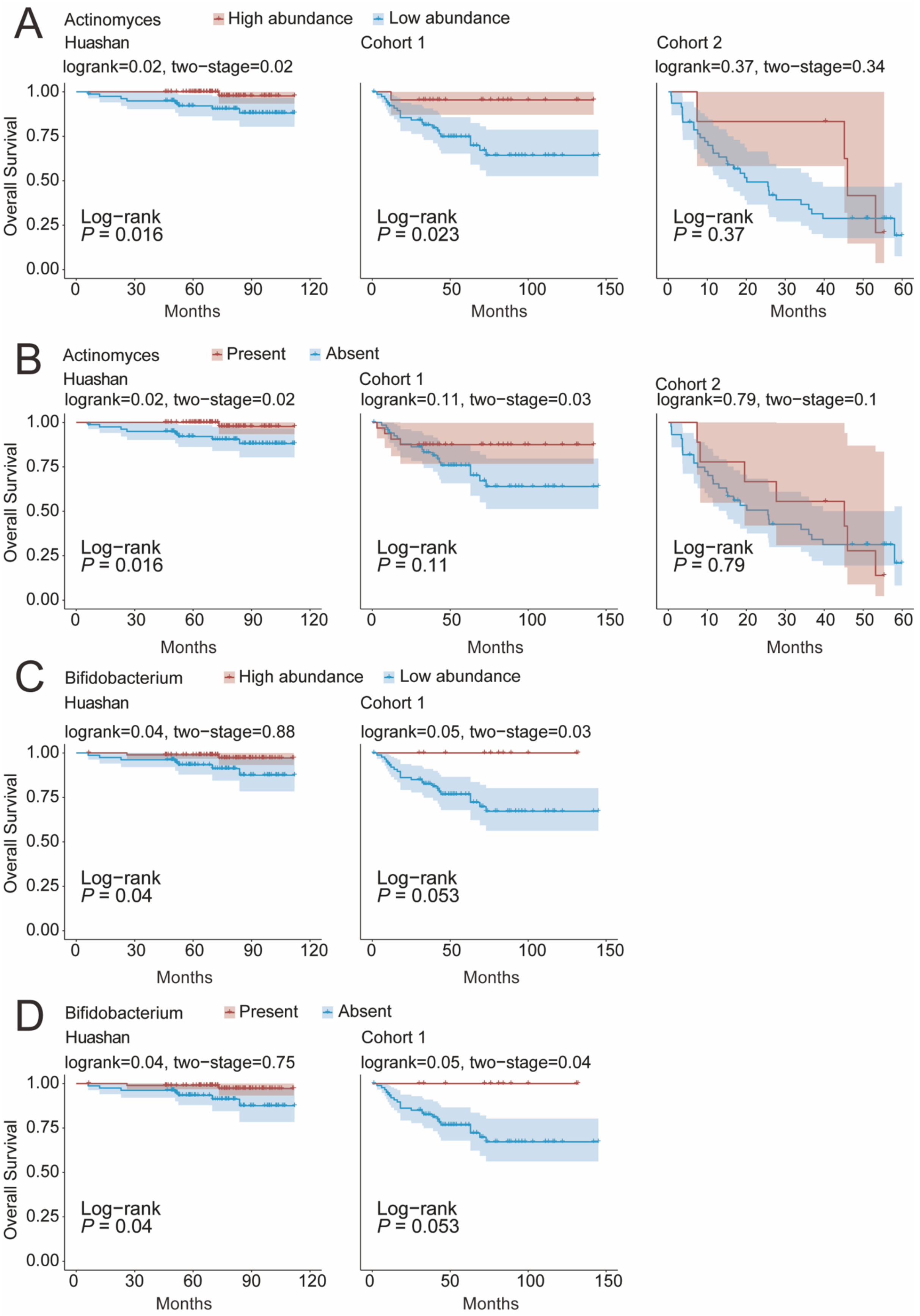

**Figure S13. Association between specific genus and overall survival**

Kaplan–Meier curves showing the overall survival probability for Huashan, Cohort 1, and Cohort 2 grouped by (A) high/low CPM abundance and (B) present/absent of *Actinomyces*. Kaplan–Meier curves showing the overall survival probability for Huashan, Cohort 1, and Cohort 2 grouped by (C) high/low CPM abundance and (D) present/absent of *Bifidobacterium*. *P* values were calculated using unadjusted Log-Rank test and Two-Stage test.

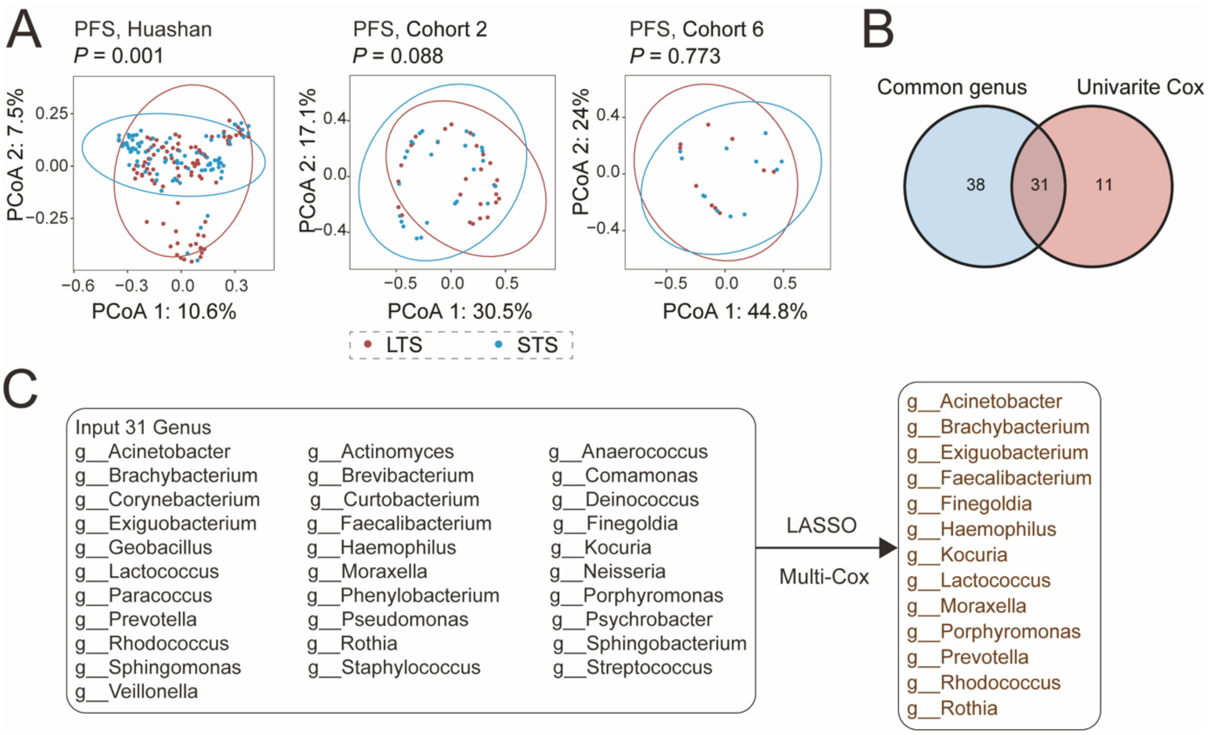

**Figure S14. Construction of microbial risk score for progression-free survival in ccRCC**

(A) Principal coordinate analysis (PCoA) for patients with long-term survival (LTS) and short-term survival (STS) of PFS in Huashan, Cohort 2 and Cohort 6, based on the Bray–Curtis dissimilarity. The *P* values were tested by Permutational multivariate analysis of variance (PERMANOVA). (B) Venn plot showed the intersections between common genera and the genera with consistent HR in univariate cox for PFS in the Huashan, Cohort 2 and Cohort 6. (C) The diagram presented the input 31 genera for the first step: LASSO. Then the selected features by LASSO were subjected to construct a Cox model. Finally, we got a microbial community used for calculating a risk score for PFS and each feature contributed positively or negatively to the score measured by the index “coef”. The genera with coef > 0 were classified as risk genera and those with coef < 0 were classified as protective genera.

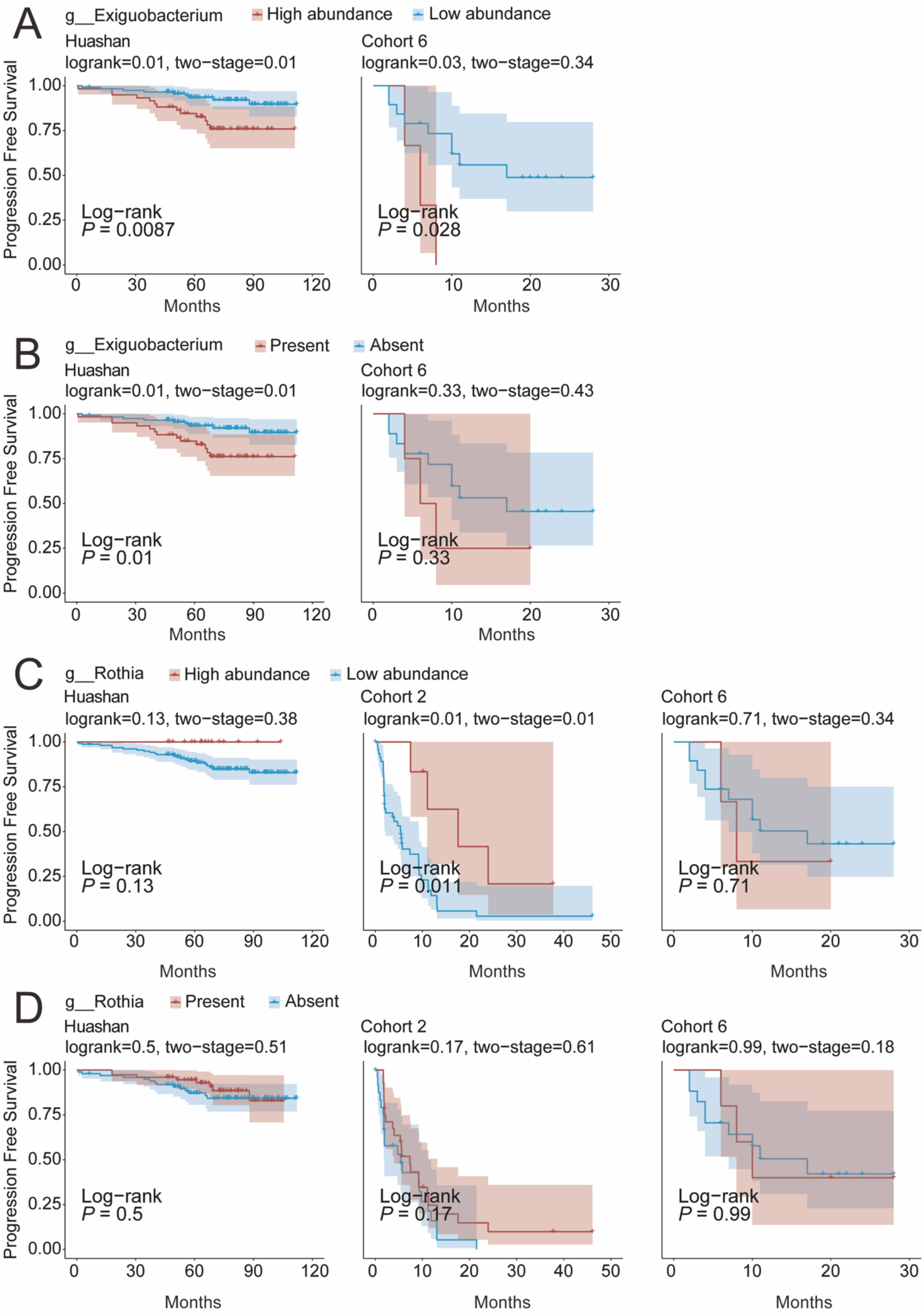

**Figure S15. Association between particular genus and progression-free survival**

Kaplan–Meier curves showing the progression-free survival probability for Huashan and Cohort 6 grouped by (A) high/low CPM abundance and (B) present/absent of *Actinomyces*. Kaplan–Meier curves showing the progression-free survival probability for Huashan, Cohort 2, and Cohort 6 grouped by (C) high/low CPM abundance and (D) present/absent of *Bifidobacterium*. *P* values were calculated using unadjusted Log-Rank test and Two-Stage test.

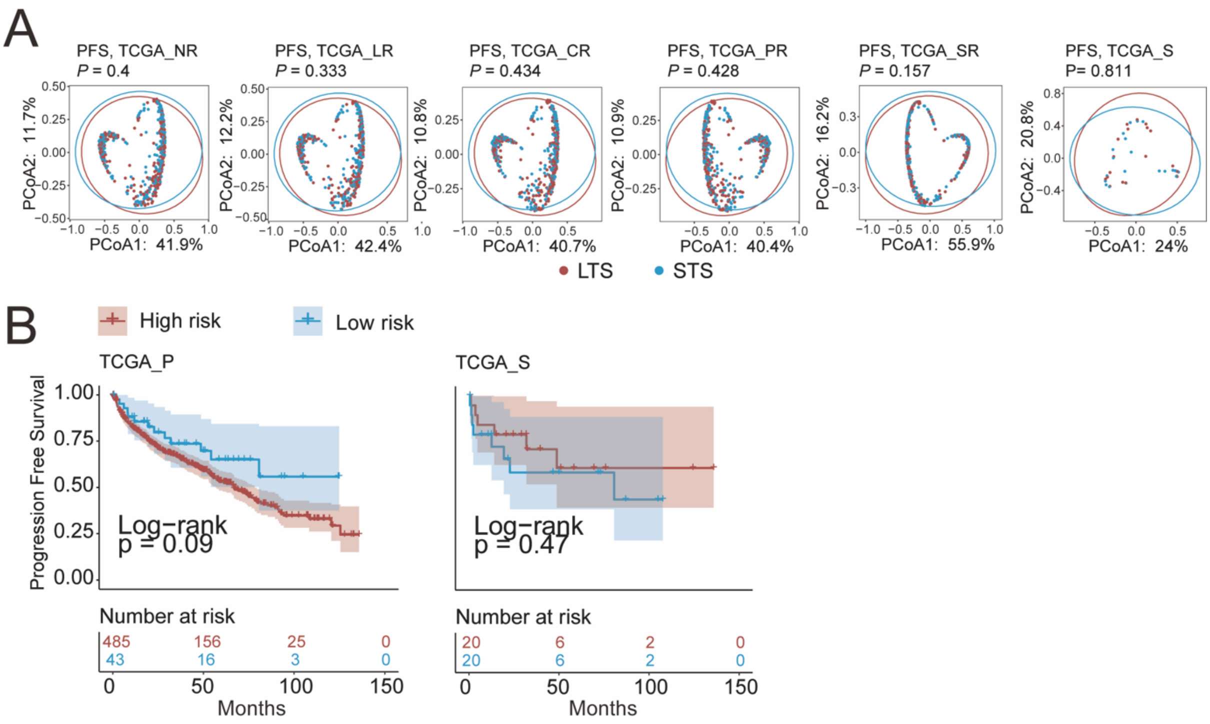

**Figure S16. Microbial risk score for PFS in TCGA**

(A) Principal coordinate analysis (PCoA) for patients with long-term survival (LTS) and short-term survival (STS) of PFS in TCGA_P(NR), TCGA_P(LR), TCGA_P(CR), TCGA_P(PR), TCGA_P(SR), and TCGA_S, based on the Bray–Curtis dissimilarity. (B) Kaplan–Meier curves showing the progression-free survival probability for TCGA_P and TCGA_S stratified by risk score. P values were calculated using unadjusted Log-Rank test.

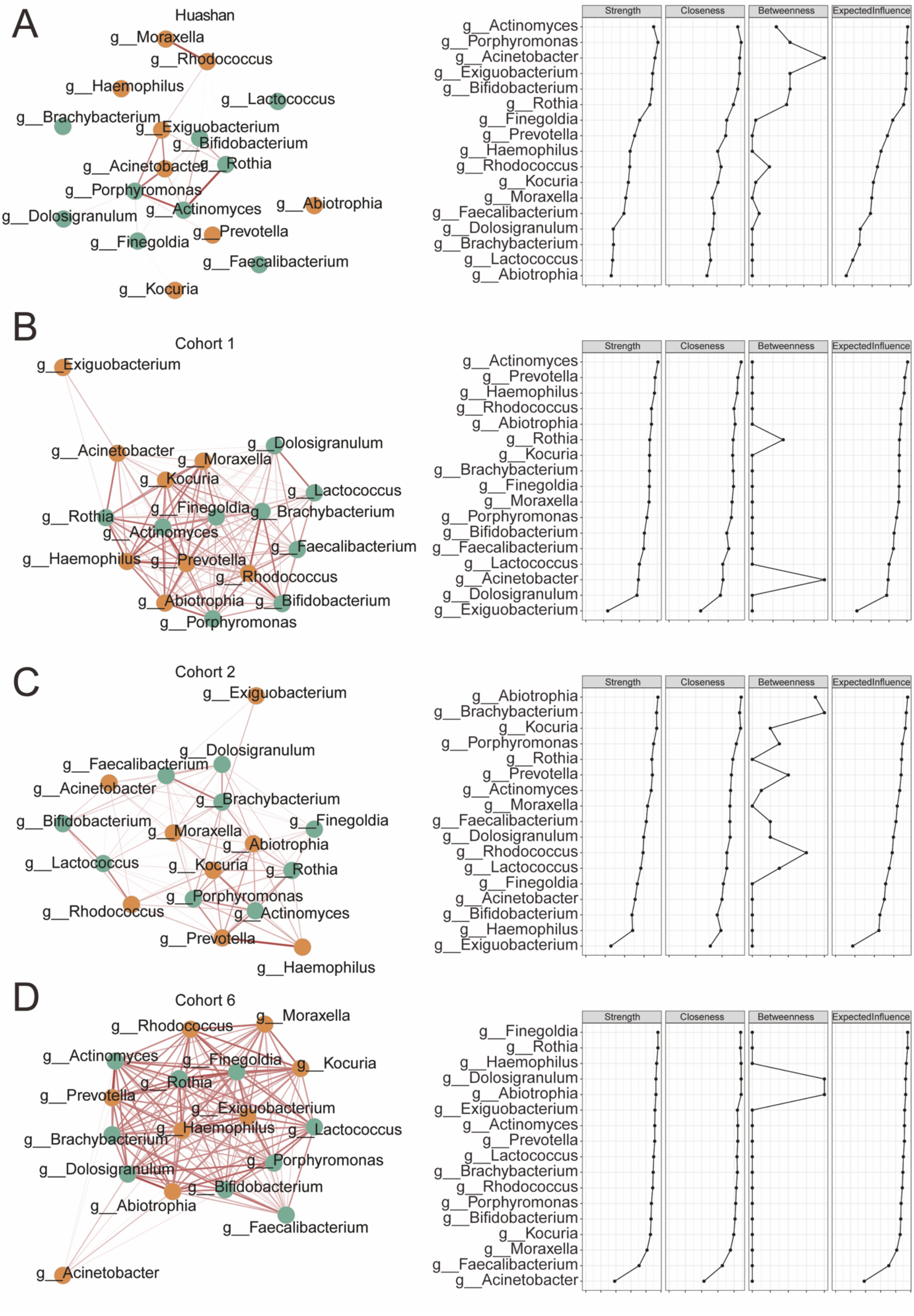

**Figure S17. Network of intratumor bacterial genera**

(A-D) The network analysis showing co-occurrence pattern of the 20 prognosis-related genera. The nodes colored yellow represented the risk genera while those colored green indicated the protective genera. A connection represents an acceptable ($r > 0.3$) and significant ($P < 0.05$) correlation. A genus (node) with great number of connections were considered as hub genus. We used four index including “Strength”, “closeness”, “Betweenness”, “ExpectedInfluence” to quantize the importance of genus.

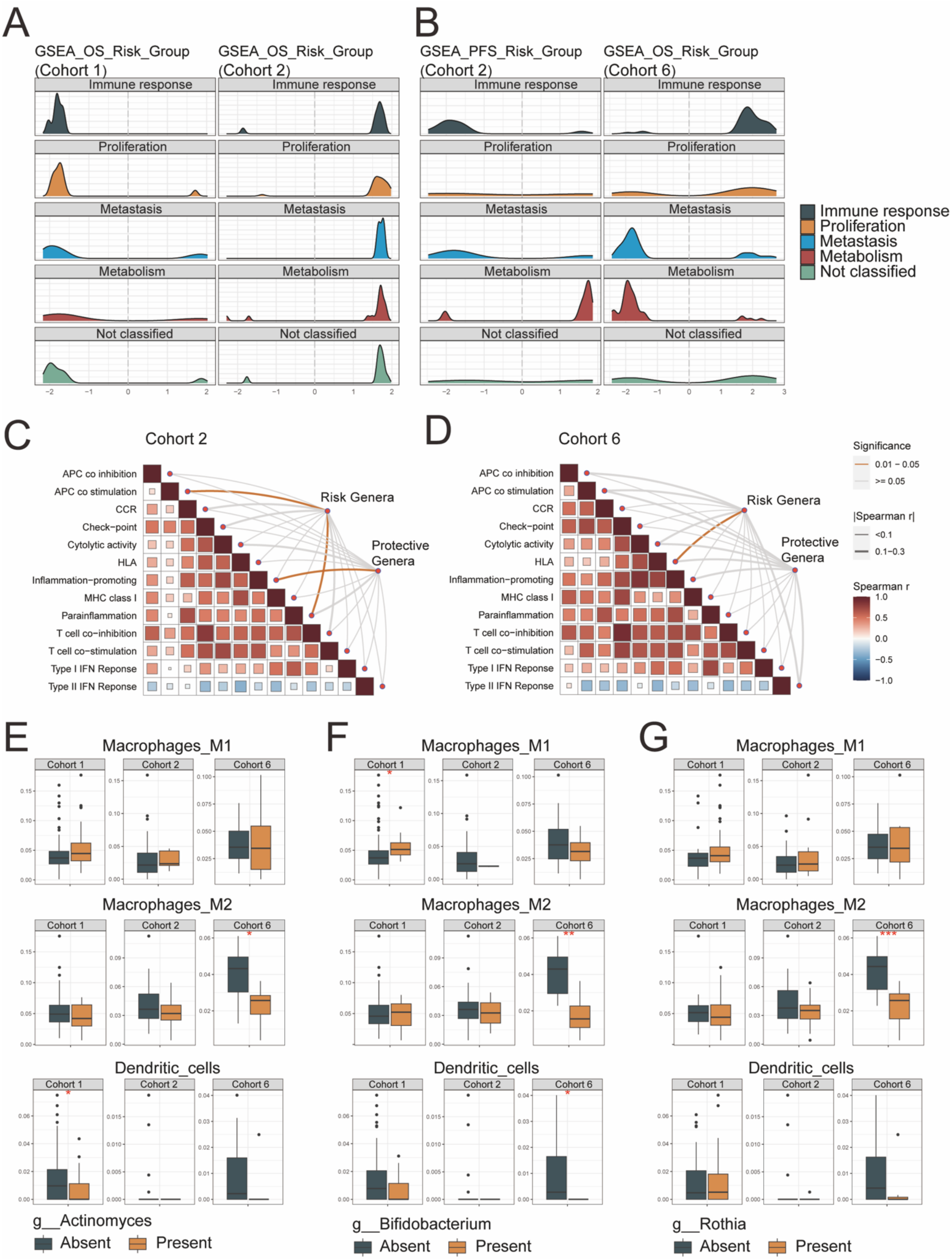

**Figure S18. Prognosis-related genera and immune**

The density curves represent the pathway distribution of the knowledge-based annotation of 4 main types: immune response, proliferation, metastasis, metabolism that were significantly enriched between the two stratified groups using gene set enrichment analysis. The horizontal axis indicated the NES of the GSEA result. The stratification was the same as the previous result, that is (A) overall survival-related risk group in Cohort 1 and 2, and (B) progression-free survival-related risk group in Cohort 2 and 6. (C-D) The result of mantel test showing the interaction between genera community and potential immune function in Cohort 2 and Cohort 6. Spearman method was used. The thickness of the curve indicated the absolute value of the spearman rho, and the significant connection was yellow colored. Each block represented the correlation among the immune functions, and a redder color meant a greater rho. Box plot exhibiting the level of M1 macrophage polarization, M2 macrophage polarization, and dendritic cells in Cohort 1, 2, 6 respectively in the presence or absence of (E) *Actinomyces*, (F) *Rothia*, (G) *Bifidobacterium*. Wilcoxon Test was used for comparing the relative abundance between tumor and normal.

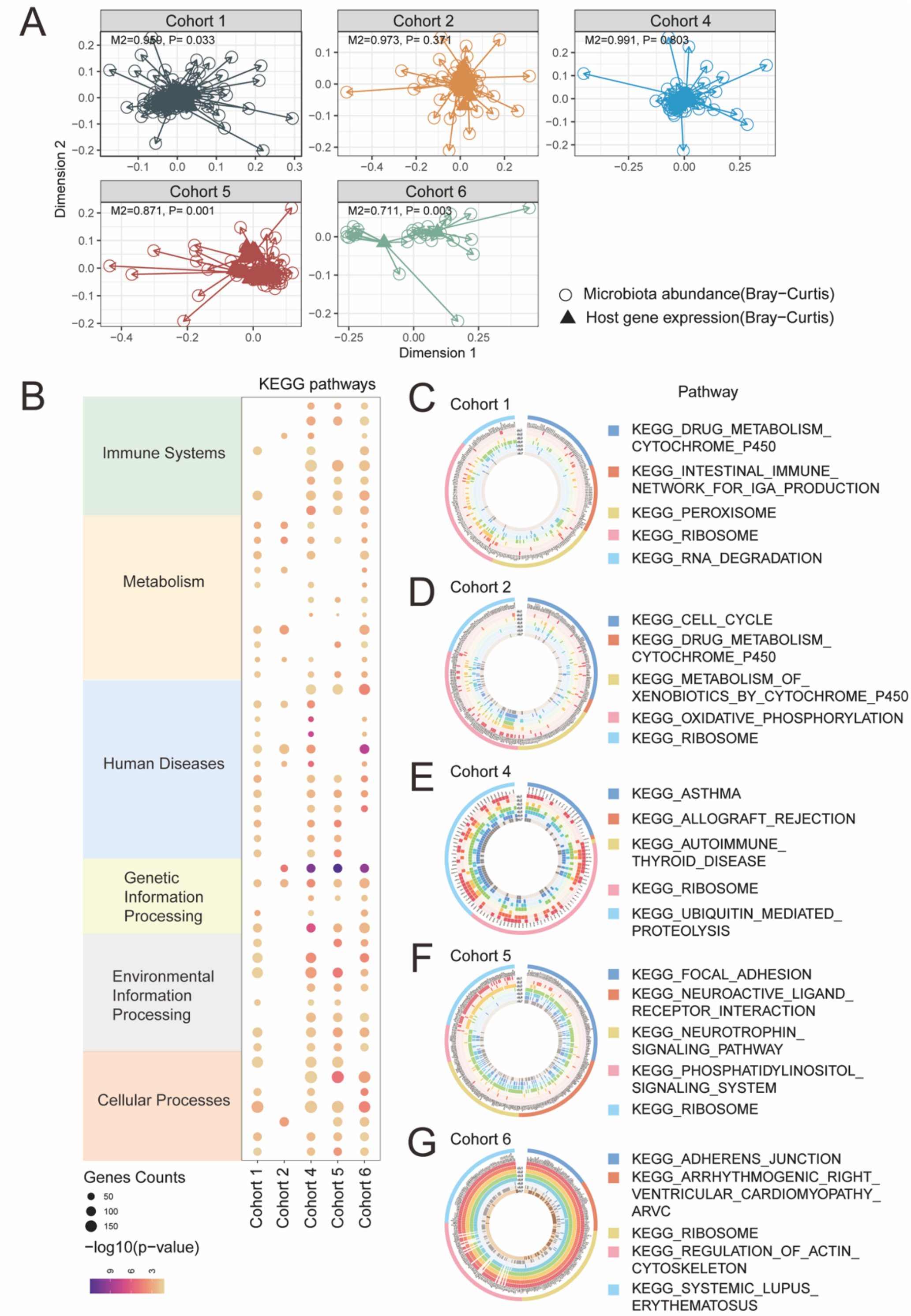

**Figure S19. Interaction between intratumor bacterial genus and host genes**

(A) Procrustes analysis showing overall association between variation in host gene expression and intratumor microbiome composition in Cohort 1,2,4,5,6. Bray-Curtis distance was used for host gene expression data (triangles) and intratumor microbiome data (circles). (B) Dot plot showing the result of KEGG pathways enriched in the genes related to the microbial communities and then the pathways were categorized into six types: immune systems, metabolism, human disease, genetic information processing, environmental information processing, and cellular processes. The size of dot represented the number hit in the pathway.

(C-G) Circular plot showing the top 5 enriched pathways in each cohort. The correlation between genes involved in the pathway and specific genus were shown in each block and were darkly colored when *P* value reached significance.
